# Therapeutic Efficacy and Safety of Deep Brain Stimulation for Multiple Sclerosis Related-Tremor: A Systematic Review and Meta-Analysis

**DOI:** 10.64898/2026.03.22.26349017

**Authors:** Farzan Fahim, MohammadAmin Farajzadeh, Seyyed Mohammad Hosseini Marvast, Mahsa Faramin Lashkarian, Ava Khalili Dehkordi, Parastoo Sangtarashha, Reihane Qahremani, Hadis Khodadadi, Mahdieh Pourabdollah, Taha Mahdian, Setareh Parsakian, MohammadReza Toghyani, Sayeh Oveisi, Guive Sharifi, Alireza Zali, Farbod Tabasi Kakhki, AmirMahdi Mojtahedzadeh

## Abstract

**Objective:** To systematically evaluate the efficacy and safety of Deep Brain Stimulation (DBS) for the management of disabling tremor in patients with Multiple Sclerosis (MS) by synthesizing data from available clinical studies.

**Methods:** This systematic review and meta-analysis followed PRISMA 2020 guidelines and was registered with PROSPERO (CRD420261347426). A comprehensive search of PubMed, Scopus, Web of Science, and Embase was performed from database inception until December 2025 with no time or language limitation. A pre-post meta-analysis design was used to estimate the pooled effect size using the Standardized Mean Change (SMC) between baseline and follow up tremor severity. Because most included studies were single arm cohorts and clinical heterogeneity was anticipated, a random effects model using the Restricted Maximum Likelihood (REML) estimator with the Hartung-Knapp adjustment was applied. Safety outcomes including hardware complications and postoperative infections were pooled using random effects meta-analysis of proportions.

**Results:** Thirteen studies including 131 patients met the eligibility criteria. Eight studies with adequate outcome data were included in the pooled efficacy analysis. DBS was associated with a significant reduction in tremor severity with an overall pooled SMC of 1.42 (95% CI 1.07 to 1.77). Statistical heterogeneity was minimal (I^2^ = 0.0%, p = 0.6300), although this finding should be interpreted cautiously given the limited number of studies and clinical variability in surgical targets, most commonly the ventral intermediate nucleus (VIM), and follow up duration ranging from months to more than 20 years. The pooled incidence of postoperative infection was approximately 7% with substantial heterogeneity across studies (I^2^ = 74.1%). The most frequently reported adverse events were stimulation related effects such as dysarthria and disequilibrium, which were generally reversible after adjustment of stimulation parameters. Overall methodological quality of included studies was predominantly moderate.

**Conclusion:** Deep brain stimulation may provide meaningful tremor reduction in selected patients with disabling and medication refractory MS tremor, with a large pooled treatment effect (SMC = 1.42). Although complications such as postoperative infection (approximately 7%) and transient stimulation related adverse effects can occur, these events appear manageable in most cases. However, the current evidence base remains limited by small sample sizes, heterogeneous study designs, and variability in surgical targets and outcome reporting. Larger prospective studies with standardized tremor outcome measures and consistent reporting of safety outcomes are needed to better define the long term efficacy and optimal clinical role of DBS in patients with MS related tremor.

## Introduction

Multiple sclerosis (MS) is a chronic immune mediated disease of the central nervous system including inflammatory demyelination and progressive neurodegeneration. It is one of the leading causes of neurological disability among mostly young adults, approximately 1.9 million patients worldwide, with a steadily rising prevalence over the past three decades (about 24 cases per 100,000 population). The highest rates occur in North America and Western Europe, particularly in Sweden, Canada, and Norway, whereas incidence in Southern regions remains considerably lower (1, 2). In addition to sensory, visual, and motor deficits, movement disorders are increasingly recognized(3). Tremor is one of the most movement disabling symptoms and can markedly affect functional independence and quality of life(4). Patients with MS related tremor often are unable to do basic daily activities such as eating, writing, and personal care, reflecting the important burden of this complication(5). Probably, tremor in MS arises from disruption of the cerebello thalamo cortical network. Demyelination involving the cerebellum, the superior cerebellar peduncle, and thalamic relay nuclei may disrupt normal motor circuit activity and produce postural and intention tremor(6). Neurophysiological studies have also demonstrated abnormalities in cerebellar output and sensorimotor integration in affected patients(7); also, diffusion tensor imaging (DTI) studies have demonstrated structural alterations in white matter pathways connecting cerebellar and thalamocortical regions which support the concept that tremor results from dysfunction within distributed motor networks(6).

Treatment of MS associated tremor remains challenging. Pharmacological therapies including anticonvulsants, benzodiazepines, and other symptomatic medications are often restricted because of limited benefit, and many patients continue to experience disabling tremor despite medical therapy(8). This limitation has led to increasing efforts in surgical approaches targeting some parts of impaired motor circuits. Deep brain stimulation (DBS) is a neuromodulatory procedure that produces controlled electrical stimulatory or inhibitory pulses to specific brain areas (including pathways or nuclei) to modulate abnormal neural activity. Some efforts established DBS as a procedure with some therapeutic effect in several movement disorders, particularly Parkinson’s disease, where it effectively reduces tremor and improves motor symptoms(9). DBS also has been explored in other neurological conditions, including Alzheimer’s disease, where stimulation of the fornix has shown potential therapeutic effects(10).

More recently, DBS has been applied to patients with multiple sclerosis and tremor, most commonly targeting the ventral intermediate nucleus of the thalamus or nearby cerebellothalamic pathways. Several observational studies have reported tremor improvement following DBS in selected patients(5, 11–13). However, the current evidence remains heterogeneous. Many studies include small sample sizes, variable patient selection, differences in stimulation targets and outcome measures, and inconsistent reporting of complications(11–14). Moreover, although tremor outcomes are frequently reported, safety, particularly device related complications such as infection, have not been systematically synthesized.

Therefore, this systematic review and meta-analysis aimed to evaluate the available evidence on deep brain stimulation for tremor reduction as primary outcome and safety of the procedure as secondary outcome in patients with multiple sclerosis.

## METHODS

### Protocol and reporting guideline

This systematic review and meta-analysis was conducted according to the Preferred Reporting Items for Systematic Reviews and Meta-Analyses (PRISMA2020) guideline(15). Also, PRISMA checklist has been provided in supplementory file 1. The study protocol was prospectively registered in the International Prospective Register of Systematic Reviews (PROSPERO)(16) under the registration number CRD420261347426. The complete protocol detailing the study rationale, eligibility criteria, and analytical plan is provided in Supplementary File 2.

### Search strategy

A comprehensive and systematic search strategy was developed by (FF an MAF) to identify studies evaluating the role of deep brain stimulation (DBS) in patients with multiple sclerosis (MS) presenting with tremor. Four electronic databases were searched: PubMed, Scopus, Web of Science, and Embase.The search strategy was designed using a combination of controlled vocabulary terms and free-text keywords. Synonyms and related concepts were identified through Medical Subject Headings (MeSH) entry terms and database indexing systems to ensure a sensitive search capturing all relevant literature. Search terms were structured around three main conceptual domains: multiple sclerosis, deep brain stimulation or neuromodulation techniques, and tremor. No language or publication date restrictions were applied. The search was conducted from database inception until December 2025. Studies published in languages other than English were not excluded; instead, such articles were translated into English using high-accuracy translation tools before entering the screening process. The complete search strategies used for all databases are provided in Supplementary File 3. The exact search syntax used in PubMed is presented below:

(“multiple sclerosis” OR “MS” OR “demyelinatingdisease*” OR “demyelination disorder*”) AND (“deep brainstimulation” OR DBS OR “neuromodulation” OR “brainstimulation” OR “stereotactic stimulation” OR “implantableneurostimulator” OR “thalamic stimulation” OR “VIMstimulation” OR “ventral intermediate nucleus stimulation”) AND (tremor OR “intention tremor” OR “postural tremor” OR “action tremor” OR “MS tremor” OR “cerebellartremor”)

### Eligibility criteria

Study selection was performed according to predefined eligibility criteria structured around the PICO framework. Eligible studies included patients diagnosed with multiple sclerosis of any subtype who experienced tremor attributable to MS. The intervention of interest was deep brain stimulation performed for tremor management, regardless of the intracranial target nucleus used for stimulation. The comparator consisted of patients who did not undergo DBS treatment or who received alternative therapeutic interventions. The primary outcome of interest was improvement or reduction in tremor severity associated with multiple sclerosis. Secondary outcomes included adverse events related to the procedure, postoperative infections, surgical complications. Eligible study designs included clinical trials and cohort studies, regardless of whether they were prospective or retrospective. Studies were excluded if they involved patients without multiple sclerosis, evaluated interventions other than deep brain stimulation, or failed to report the primary outcome related to tremor improvement. Case reports, case series, conference abstracts, review articles, and animal studies were also excluded from the analysis.

### Study selection and screening process

All retrieved records were imported into Covidence for reference management and duplicate removal. Title and abstract screening was conducted independently by two reviewers (AK and MP). During this stage, each reviewer maintained a separate Excel file documenting excluded studies along with the study title, DOI, and the specific reason for exclusion. The complete documentation of excluded studies during title and abstract screening is provided in Supplementary File 4. Full-text screening was also performed independently by two reviewers (PS and MH). Each reviewer generated two separate Excel files: one documenting the studies included for final analysis and another documenting studies excluded after full-text assessment. The file containing included studies recorded the study title, DOI, reported outcomes, type of control group, and study design. The file containing excluded studies recorded the study title, DOI, country of origin, first author, and the reason for exclusion. The senior writer (FF) examined and fixed their inconsistencies. All documentation related to the full-text screening process is available in Supplementary File 5.

### Data extraction

A standardized data extraction sheet was developed by the senior author(FF) prior to the start of the data extraction process. Reviewers were trained in the use of the extraction form to ensure consistency and methodological rigor. Two reviewers independently (MF and MH) extracted data from all included studies. The extraction information regarding study characteristics, patient demographics, disease-specific variables, intervention details, outcome measurements, adverse events, and methodological considerations. Study-level variables included the study title, author and year of publication, DOI, funding source, geographic region, study design, center type, period of data collection, inclusion and exclusion criteria, follow-up duration, ethical approval status, and declarations of funding or conflicts of interest. Population-level variables included the number of cases and controls, mean age in intervention and control groups, and sex distribution. Disease-specific variables related to multiple sclerosis included MS subtype, patient age, disease duration, baseline tremor characteristics, tremor measurement instruments, Expanded Disability Status Scale (EDSS) at baseline, EDSS range, presence of cerebellar signs, baseline cognitive impairment, tremor type, tremor distribution, tremor duration prior to DBS implantation, previous medical therapies for tremor, and definitions of medication-refractory tremor when reported. Detailed intervention-related variables were also extracted, including DBS target nucleus, anatomical targeting method, stimulation laterality, number of active contacts, number of implanted electrodes, stimulation amplitude or voltage, stimulation frequency, pulse width, stimulation configuration (monopolar versus bipolar), time from MS diagnosis to DBS implantation, time from tremor onset to DBS implantation, number of programming sessions, time to optimal stimulation parameters, and whether stimulation parameters were modified during follow-up. Outcome variables included tremor severity scores at baseline and follow-up with corresponding standard deviations for both intervention and control groups when available. Safety outcomes were also extracted, including surgery-related complications, stimulation-related adverse events, neurological worsening, infections, and intracranial hemorrhage. The senior writer (FF) examined and provided final extraction excel sheet. The complete data extraction sheet is provided in Supplementary File 6.

### Risk of bias assessment

The methodological quality of the included studies was independently evaluated by two reviewers (TM and HK) using the Joanna Briggs Institute (JBI) critical appraisal tools(17). Separate checklists were applied according to study design. For randomized clinical trials, the JBI checklist evaluates methodological domains including randomization procedures(18), allocation concealment, similarity of groups at baseline, blinding of participants, blinding of those delivering the intervention, blinding of outcome assessors, completeness of follow-up, consistency and reliability of outcome measurement, and appropriateness of statistical analysis. For cohort studies(19), the JBI checklist assesses whether the two groups were similar and recruited from the same population, whether exposures were measured in a valid and reliable way, whether confounding factors were identified and addressed, whether outcomes were measured validly and reliably, whether follow-up was sufficiently long for outcomes to occur, whether follow-up was complete, and whether appropriate statistical analysis was used. Each reviewer completed the risk-of-bias checklist independently. Disagreements were resolved by The senior writer (FF). Completed risk-of-bias assessment form are presented in Supplementary File 7.

### Statistical Analysis

All quantitative analyses were conducted using R statistical software (R Foundation for Statistical Computing, Vienna, Austria; version 4.5.1) with the meta and metafor package by senior author (FF). The primary objective of the meta-analysis was to estimate the overall change in tremor severity following deep brain stimulation (DBS) in patients with multiple sclerosis (MS). Because the included studies were predominantly single-arm cohorts without control groups, a pre–post meta-analysis design was used. The treatment effect was calculated as the standardized mean change (SMC) between preoperative baseline tremor severity and postoperative tremor severity at the latest reported follow-up.

### Data conversion procedures

In several studies, complete summary statistics were not reported in the required format for meta-analysis. For example, some studies reported tremor scores graphically or presented dispersion measures as standard error of the mean (SEM), median with range, or other incomplete formats. In such cases, standard statistical conversion methods were applied to estimate the required mean and standard deviation values. When standard error (SEM) was reported instead of standard deviation (SD), SD was calculated using the formula: SD = SEM × √n. where n represents the study sample size.

When data were reported as median and range, mean and standard deviation were estimated using the method described by Hozo et al(20). This approach estimates the sample mean and variance from the reported median, minimum, maximum, and sample size. were applied:

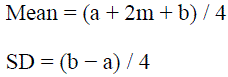

where a is the minimum value, b is the maximum value, and m is the median. These conversion approaches are widely used in meta-analysis when full summary statistics are unavailable and allow inclusion of studies that otherwise could not contribute to pooled quantitative synthesis.

### Effect size calculation

Effect sizes were calculated using the standardized mean change with raw score standardization (SMCR), which accounts for within-subject correlation between pre– and postoperative measurements. The SMC was computed using the escalc function in the metafor package. Because the correlation between baseline and follow-up measurements was not reported in the included studies, a correlation coefficient of r = 0.5 was assumed, which is a commonly applied assumption in pre–post meta-analyses when the true correlation is unknown.

#### Random-effects Meta-analysis (REML; Hartung–Knapp)

To synthesize the evidence, a random-effects meta-analysis was employed. The random-effects model was chosen a priori due to anticipated clinical heterogeneity among studies, including differences in surgical targets, types and comparisons of tremor rating scales, durations of follow-up, and demographic characteristics of patient populations. Estimates of between-study variance were obtained using the restricted maximum likelihood (REML) method. Additionally, to enhance the coverage of uncertainty estimates in the context of a limited number of studies, the Hartung–Knapp (HK) adjustment was employed. This adjustment is particularly beneficial in mitigating the bias that can arise from small sample sizes, ensuring more reliable confidence intervals.

#### Effect Size and Display in Forest Plot

The effect size in this analysis was reported using the Standardized Mean Change (SMC). SMC facilitates the comparison of intervention effects across studies that may employ different measurement scales or units. In the forest plot, the effects of individual studies were displayed alongside their corresponding 95% confidence intervals, and subsequently, a pooled effect estimate was calculated based on the random-effects model.

#### Evaluation of Heterogeneity

Statistical heterogeneity among studies was assessed using the I² statistic and τ² (between-study variance). Specifically, I² quantifies the percentage of total variation across studies that is attributable to true heterogeneity rather than chance. τ², on the other hand, provides an estimate of the variance among the studies applying the random-effects model. The corresponding p-value for the heterogeneity test was reported to inform whether the observed heterogeneity was statistically significant.

#### Subgroup Analysis Based on Data Acquisition Method

To investigate whether the data extraction methods could influence the pooled effect estimate, a subgroup analysis was performed based on the method of data acquisition. The studies were categorized into two groups:

1. Direct: Studies that provided outcomes of tremor in a directly extractable format (mean ± SD reported).
2. Converted: Studies that required statistical conversion of summary statistics to derive the necessary data.

#### Publication bias assessment

Potential small-study effects and publication bias were explored using funnel plot visualization, in which the standardized mean change was plotted against the corresponding standard error for each study. Because the total number of included studies was fewer than ten, formal statistical tests for funnel plot asymmetry were interpreted cautiously.

#### Sensitivity analysis

A leave-one-out sensitivity analysis was conducted to evaluate the robustness of the pooled effect estimate. In this procedure, the meta-analysis was repeated iteratively after removing one study at a time, allowing assessment of whether any single study exerted a disproportionate influence on the overall effect size.

#### Meta-regression

Exploratory meta-regression analyses were performed to investigate whether selected study-level characteristics were associated with variations in effect size. The moderators evaluated included follow-up duration, study sample size, and data extraction method (direct versus converted data). Meta-regression models were fitted using mixed-effects models with REML estimation. Given the limited number of included studies, meta-regression analyses were considered exploratory and interpreted cautiously.

#### Safety outcomes

In addition to efficacy outcome, safety outcomes were analyzed. Separate random-effects meta-analyses of proportions were conducted to estimate the pooled rate of surgical complications and postoperative infections associated with DBS in patients with MS. Proportions and their 95% confidence intervals were calculated for each study, and pooled estimates were generated using random-effects models. Heterogeneity was assessed using I² and τ² statistics.

## Results

### Study selection and Characteristics

In total, 1,012 records were identified through database searching (Embase n = 301, Scopus n = 262, Web of Science n = 261, PubMed n = 188). After removal of 470 duplicates, 542 unique records remained for title and abstract screening. Of these, 464 were excluded as clearly irrelevant. Seventy-eight full-text articles were sought for retrieval and all were successfully obtained. After full-text assessment, 65 articles were excluded for the following reasons: conference abstract only (n = 25), case report (n = 6), case series (n = 17), protocol only (n = 3), not a relevant intervention (n = 2), not an eligible patient population (n = 6), tremor outcome not extractable (n = 3), or not addressing the prespecified scientific question (n = 3). Consequently, 13 studies (3 RCTs, 1 non-randomized trial, 6 retrospective cohorts, 3 prospective cohorts) included. The selection process is summarized in the PRISMA 2020 flow diagram (Figure 1). (table 1)

**Figure 1.**
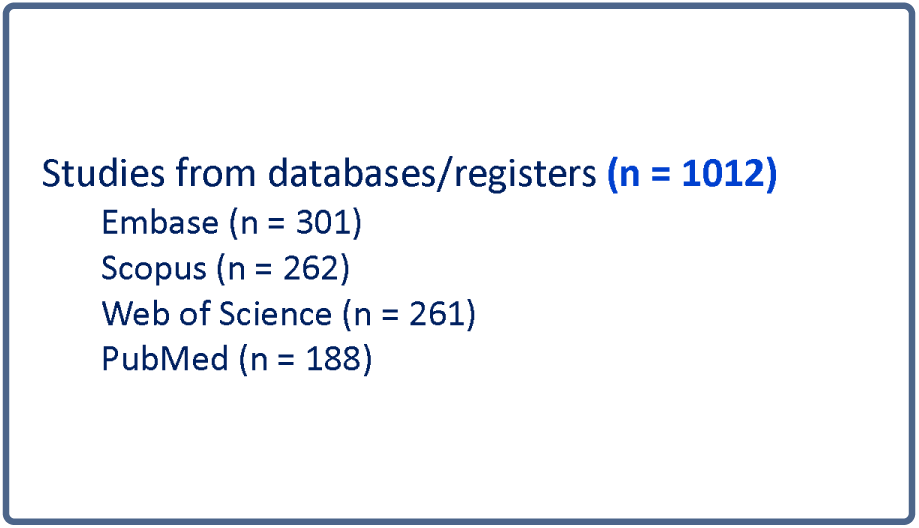
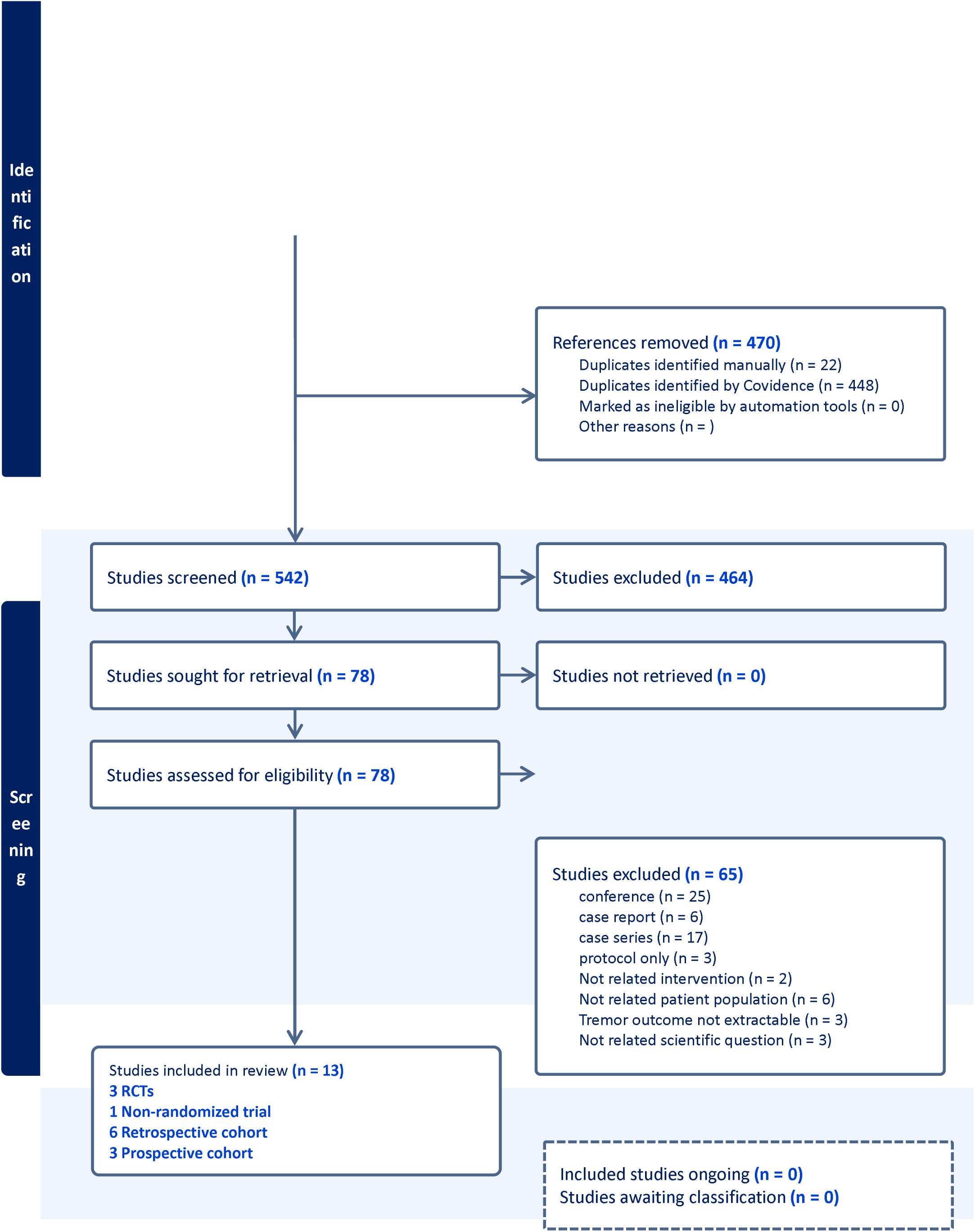
PRISMA flowchart.

**Table 1.**
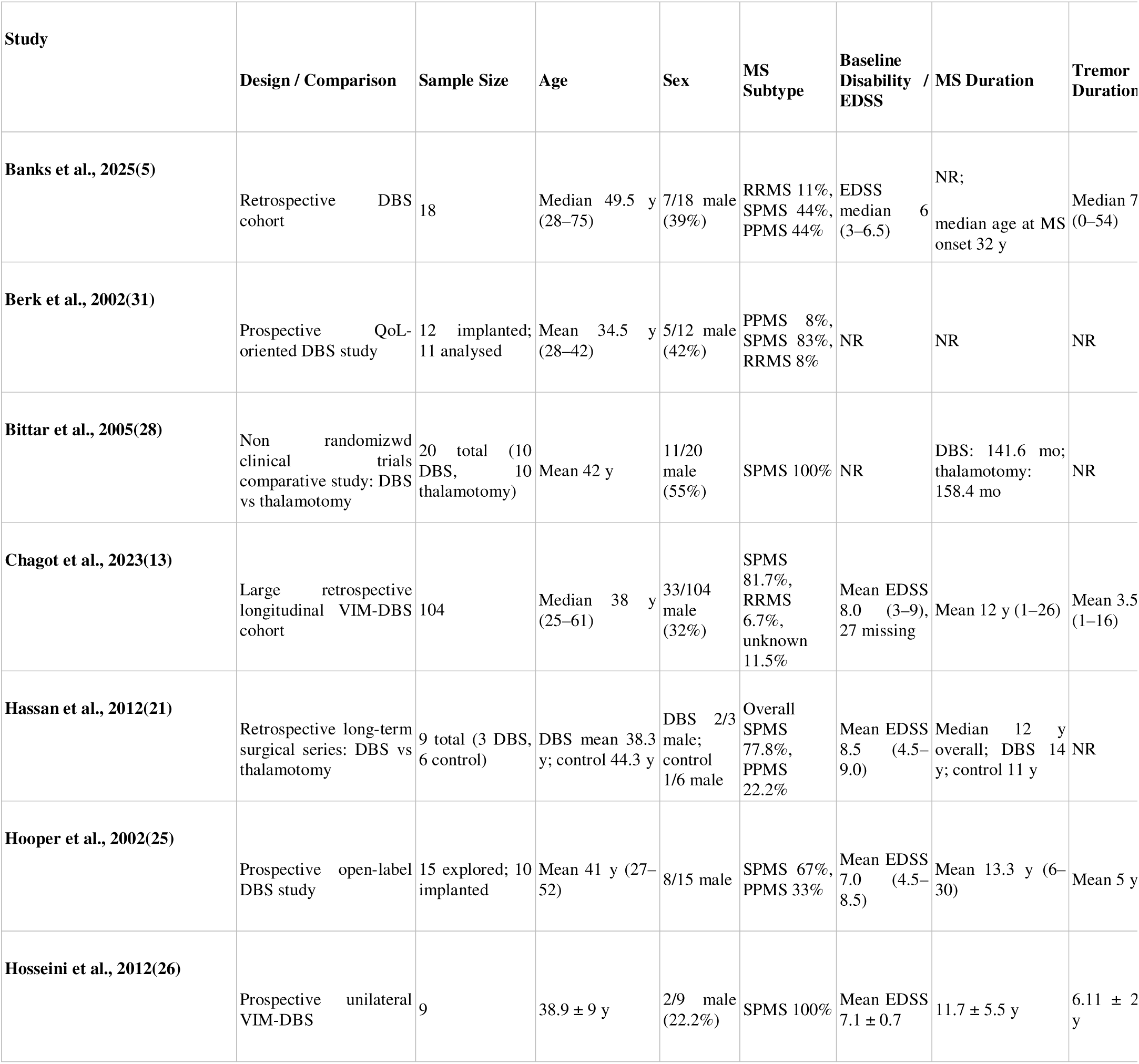
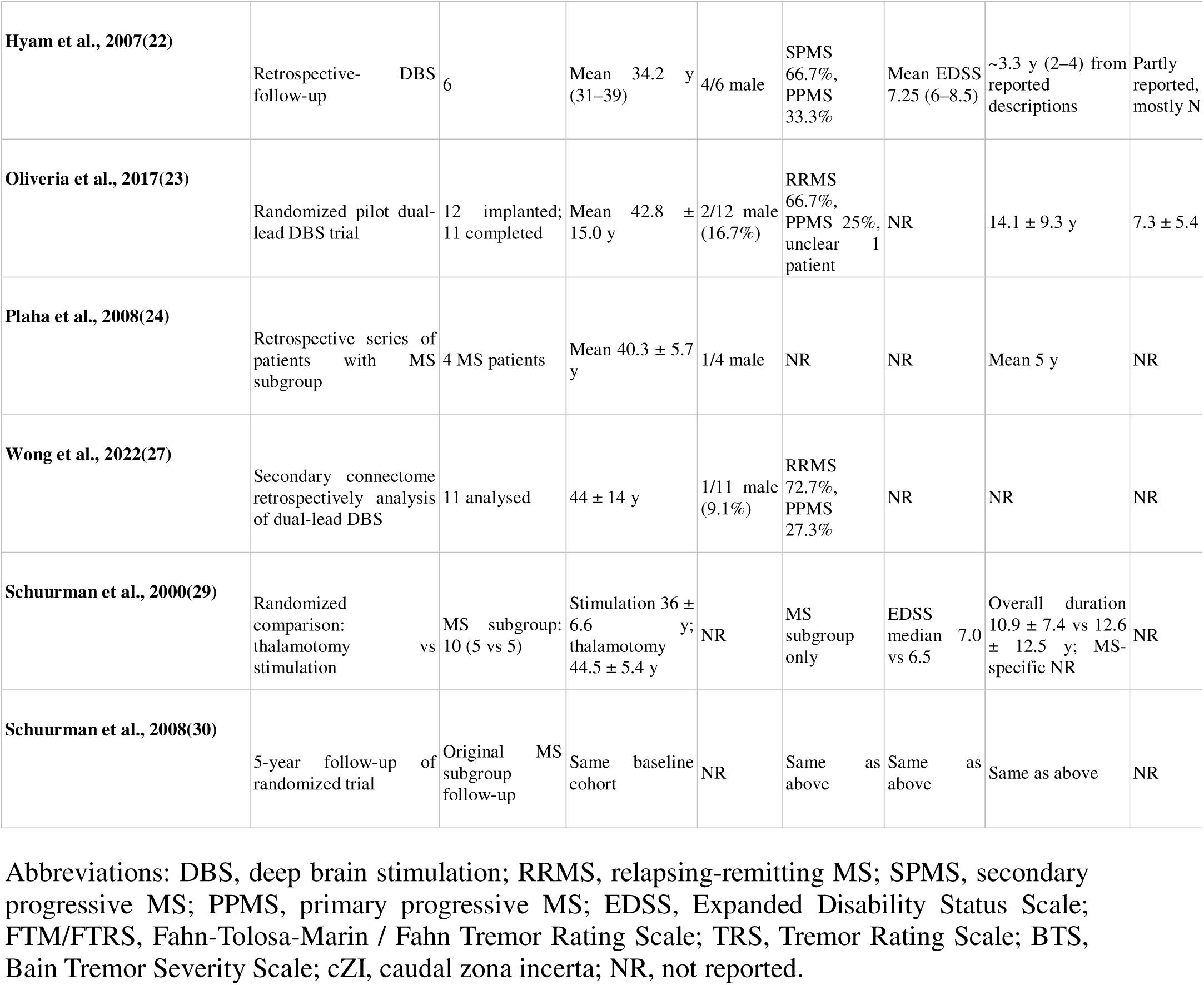
Baseline Characteristics of Included Studies.

### Patients’ characteristics

The total 131 patients included were adults (ranged in: 25-77, mean age: 42.3) with long-standing MS and disabling tremor, most frequently with progressive disease phenotypes. Sample sizes and MS subtypes varied across studies, ranging from small series of 3–12 individuals (e.g., Hassan et.al (21), Hyam et.al (22), Oliveira et.al (23) and Plaha et.al (24)) to larger cohorts such as Banks et.al(5) (18 patients) and Chagot et al.(13) (104 patients), the latter predominantly comprising secondary progressive MS. Patients were generally in the fourth to fifth decades of life, with typical MS durations of 5–15 years before surgery and similarly long tremor durations reflecting chronic, treatment-refractory symptoms. Across studies, including Banks et al.(5), Chagot et al.(13), Hooper et al.(25), Hosseini et al.(26), and Oliveira et al.(23), tremor had often persisted for several years prior to DBS, indicating substantial cumulative motor disability before intervention.

### Details of Deep brain stimulation across studies

DBS targets varied, although the ventral intermediate nucleus (VIM) was the most commonly used site across the majority of studies. Some investigations employed alternative or combined targets, including the ventralis oralis posterior (VOP), zona incerta (ZI), and caudal ZI (cZI), while Oliveira et al.(23) and Wong et al.(27) used dual thalamic leads to engage broader tremor-network pathways. Implantation laterality differed as well, with most procedures being unilateral, although Banks et al.(5), Plaha et al.(24), and Hyam et al.(22) included bilateral approaches. Follow-up durations varied: many studies reported short-term outcomes at 3–12 months, whereas others provided long-term data extending from 3 to over 20 years. This heterogeneity in targeting strategies, patient characteristics, and follow-up lengths contributes to variability across studies and complicates definitive conclusions regarding the long-term durability of DBS effects in MS tremor.

### Tremor reduction; qualitive and quantitative

Clinical efficacy was systematically evaluated using standardized rating scales, predominantly the Fahn–Tolosa–Marin Tremor Rating Scale, the standard Tremor Rating Scale, and the Bain tremor scale. Across studies targeting the VIM nucleus, substantial immediate postoperative attenuation of resting, postural, and kinetic tremor was consistently documented. In the cohort reported by Hooper et al.(25), within-subject ON versus OFF stimulation comparisons demonstrated marked clinical benefit; however, these analyses were unadjusted for multivariable confounders and specific P-values were not available in the extracted data. Hosseini et al.(26) similarly utilized a single-arm ON versus OFF design for tremor scores, demonstrating clinically relevant improvement, again without provision of exact P-values in the extraction files. In comparative analyses such as the trial by Bittar et al.(28), outcomes were explicitly contrasted between thalamotomy and chronic DBS. At a mean follow-up of approximately 15 months, the DBS cohort demonstrated a 64% improvement in postural tremor and a 36% reduction in intention tremor. While thalamotomy exhibited slightly superior efficacy for intention tremor suppression (72% improvement; P < 0.05) within that specific cohort, the intrinsic adjustability and reversibility of DBS remained critical clinical considerations. The randomized trials by Schuurman et al.(29, 30) also compared VIM-DBS directly against thalamotomy using multivariable analyses; however, specific statistical outputs and P-values for the MS subgroup’s tremor reduction were not reported in the extracted dataset. Studies exploring alternative anatomical targets reported distinct efficacy profiles. Plaha et al.(24) targeted the caudal zona incerta, but formal statistical testing for the MS subgroup was omitted. Hyam et al.(22) investigated non-VIM thalamic and subthalamic regions using unadjusted t-tests and correlation analyses, yet quantitative P-values were not reported. Oliveira et al.(23) provided qualitative assessments of continuous stimulation efficacy, explicitly noting that the study was not powered to detect between-condition statistical differences. Wong et al.(27) employed a dual-lead strategy and focused their secondary analysis on connectomic associations with the percentage of tremor improvement at 6 months, identifying network connectivity metrics associated with optimal outcomes, although raw P-values for these clinical improvements were not reported. Banks et al.(5) similarly engaged in exploratory correlation analyses of tremor suppression but did not report absolute P-values for the primary tremor change outcomes. Table 3

A total of eight studies were included in the quantitative meta-analysis evaluating the effect of deep brain stimulation (DBS) on tremor severity in patients with multiple sclerosis.

#### Pooled Meta-analysis

In the analysis employing a random-effects model with the REML and Hartung–Knapp (HK) adjustment, the pooled analysis demonstrated a significant reduction in tremor severity following deep brain stimulation (DBS). The overall standardized mean change (SMC) was found to be 1.42 (95% CI 1.07 to 1.77). From a statistical heterogeneity perspective, the studies exhibited an I² of 0.0%, indicating no observed heterogeneity among the studies, alongside a τ² of < 0.0001 (p = 0.6300). This pattern indicates minimal to no statistically significant variance between studies regarding effect size, suggesting that the variability in observations can be attributed more to chance rather than substantial differences in true effects. Given the limited number of studies available for this meta-analysis, the combined application of the Hartung–Knapp adjustment alongside the REML estimator serves to enhance the reliability of uncertainty estimates. Consequently, the reported confidence intervals reflect a more cautious interpretation of the effects observed. (figure 2)

**Figure 2:**
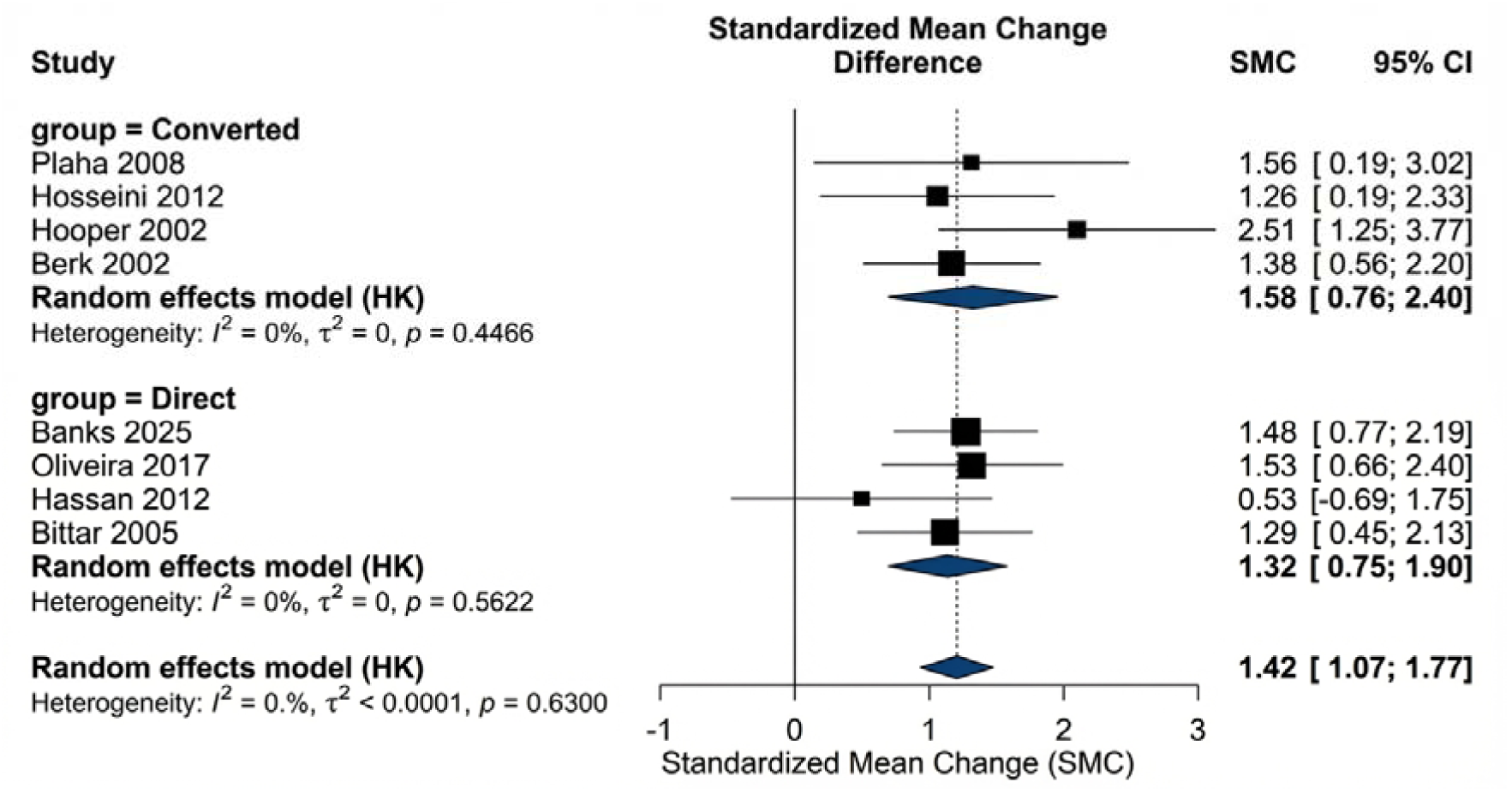
pooled and subgroup meta-analysis of tremor reduction.

#### Range of Individual Study Effects

The effect sizes across individual studies varied between 1.26 to 2.51. Among these, the largest effect size was recorded in the study by Hooper 2002, which reported an SMC of 2.51 (95% CI 1.25 to 3.77). Conversely, the smallest effect size was noted in Hosseini 2012, with an SMC of 1.26 (95% CI 0.19 to 2.33). While some confidence intervals for individual studies may appear marginal or broad, the direction of effect remained consistently in favor of improvement following DBS across all studies.

#### Subgroup Analysis

To explore the potential influence of different data extraction methods on the effect size, subgroup analyses were categorized into Converted and Direct methodologies. (figure 2)

1. Converted Subgroup (Studies Requiring Statistical Conversion): In this subgroup, the pooled effect size calculated using the random-effects model yielded an SMC of 1.58 (95% CI 0.76 to 2.40). For heterogeneity within the Converted subgroup, an I² of 0% and τ² of 0 were reported (p = 0.4466). This lack of significant heterogeneity supports the assumption of similarity across studies within this subgroup. Individual studies included in the Converted subgroup were:
  ○ Plaha(24) 2008: SMC = 1.56 [0.19; 3.02]
  ○ Hosseini(26) 2012: SMC = 1.26 [0.19; 2.33]
  ○ Hooper(25) 2002: SMC = 2.51 [1.25; 3.77]
  ○ Berk(31) 2002: SMC = 1.38 [0.56; 2.20]
2. Direct Subgroup (Studies with Direct Mean ± SD Extraction): The pooled effect size observed in the Direct subgroup was 1.32 (95% CI 0.75 to 1.90). The heterogeneity reported for this subgroup was also minimal, with an I² of 0% and τ² of 0 (p = 0.5622). The studies included within the Direct subgroup were:
  ○ Banks(5) 2025: SMC = 1.48 [0.77; 2.19]
  ○ Oliveira(23) 2017: SMC = 1.53 [0.66; 2.40]
  ○ Hassan(21) 2012: SMC = 0.53 [-0.69; 1.75]
  ○ Bittar(28) 2005: SMC = 1.29 [0.45; 2.13]

This aggregation indicates that although individual studies such as Hassan(21) 2012 demonstrate confidence intervals encompassing zero (suggesting a potentially non-significant effect), the overall pooled estimate for the Direct subgroup continued to favor improvement. Importantly, confidence intervals mainly covered the positive range, supporting the hypothesis of efficacy post-DBS.

#### Subgroup Difference Testing

To evaluate whether disparities in data extraction methods significantly impacted the effect size, a test for subgroup differences was conducted. The analysis revealed a statistic of χ² = 0.66 with df = 1, resulting in a p-value of 0.4166.

Consequently, there is insufficient statistical evidence to claim a significant difference in effect size estimates between the Converted and Direct subgroups. In summary, the direction and magnitude of the overall effect were consistently aligned across both data extraction methods, indicating that the method of data conversion does not appear to be a pivotal factor influencing the final outcome estimate.

### Quality of Life and Activities of Daily Living

The assessment of Quality of Life and Activities of Daily Living was characterized by significant methodological variability and a high degree of missing quantitative data within the final extracted dataset. While assessment tools were frequently referenced in the methodologies, final numerical scores were predominantly not reported. Berk et al.(31) monitored these domains over a 12-month period utilizing the Short Form 36 and standardized metrics, but the precise numerical values for the follow-up scores were not reported. Chagot et al.(13) captured baseline impairments specifically related to writing, dressing, and eating, but a dedicated, comprehensive questionnaire was not recorded. Hassan et al.(21) extracted a tremor-related disability scale rather than a generalized instrument, while Hooper et al.(25) assessed hand function using the Jebsen Test of Hand Function, for which quantitative outcomes were not reported. Hosseini et al.(26) utilized a functional disability scale ranging from 0 to 24, recording a baseline median functional disability score of 20, but numerical follow-up values representing post-DBS changes were not reported. Hyam et al.(22) monitored daily activities using the Barthel Index without providing exact pre– and post-operative numbers. Oliveria et al.(23) assessed functional disability exclusively via the questionnaire embedded within the Tremor Rating Scale. The trials by Schuurman et al.(29, 30). utilized the Frenchay Activities Index as a covariate within their statistical models, yet the specific numerical data for the MS subgroup at baseline and follow-up were not reported. Furthermore, in the datasets for Banks et al.(5), Bittar et al.(28), Plaha et al.(24), and Wong et al.(27), standard quantification tools for these secondary outcomes were strictly not reported, with Banks explicitly noting in their limitations that standard evaluation was severely restricted.

### Cerebellar Ataxia Cognitive Function Outcomes

The differentiation between severe MS-induced tremor and underlying cerebellar ataxia, alongside the preservation of cognitive function, emerged as critical variables across the study evaluations. Quantitative psychometric or ataxia scales were generally not recorded; instead, these variables were treated as qualitative clinical observations or strict exclusion criteria. Ataxia was frequently cited as a confounding neurological deficit that limited the apparent efficacy of DBS. Banks et al.(5)highlighted the difficulty of differentiating tremor from ataxia during patient selection, while Berk et al.(31) explicitly concluded that pre-existing ataxia is not ameliorated by DBS. The limitations in Chagot et al.(13) explicitly stated there was no data on concomitant cerebellar ataxia. Hassan et al.(21) documented the presence of cerebellar outflow tremor, and Hooper et al.(25) reported that all enrolled subjects exhibited distinct cerebellar and pyramidal signs at baseline, noting that dysmetria and ataxia remained stable and persistent post-DBS. Hosseini et al.(26) screened patients to ensure cerebellar involvement was minimal prior to surgery. Hyam et al.(22) referenced dysarthria and ataxia qualitatively within individual case descriptions. Oliveria et al.(23) imposed strict exclusion criteria for severe cerebellar dysfunction, later revealing in secondary analyses that the majority of non-responders possessed severe underlying ataxia. Plaha et al.(24) documented truncal ataxia and scanning speech in a quarter of their subjects, and Schuurman et al.(29, 30) formally categorized arm ataxia, severe gait disturbances, and balance dysfunction under adverse events rather than scoring them on a continuous severity scale.

### Cognitive Function Outcomes

Cognitive impairment was primarily utilized as a preoperative threshold for surgical exclusion rather than a longitudinally measured outcome. Chagot et al., Hosseini et al., and Schuurman et al.(13, 26, 29) explicitly excluded patients presenting with baseline cognitive impairment, defined uniformly as a Mini-Mental State Examination score of less than 24 out of 30, rendering subsequent quantitative cognitive changes not reported. Banks et al.(5) admitted to a limited cognition assessment protocol, restricting post-operative cognitive metrics to qualitative reports of speech decline. Berk et al.(31) excluded severe cognitive impairment but noted their index patient was highly cognitively impaired, though specific psychometric test results were not reported. Hooper et al.(25) acknowledged that approximately 40% of their cohort exhibited some degree of cognitive deficit at baseline, yet specific testing tools and post-operative trajectories were not reported. Hassan et al.(21) exclusively documented cognitive decline as a manifestation of long-term global MS progression. Data regarding cognition was entirely not reported for the cohorts analyzed by Bittar et al., Hyam et al., Oliveria et al., Plaha et al., and Wong et al.(22–24, 27, 28)

### Safety and adverse events

Postoperative complications were reported across nearly all included studies, with varying frequency and severity. Although most complications were mild or transient, several studies documented clinically meaningful events requiring additional surgical or medical management. Surgical complications were relatively uncommon but occasionally serious. Across pooled data from 13 studies the overall surgical complication rate was approximately 6–8%, as derived from the meta-analysis forest plot. The most frequent events included postoperative wound infection and intracranial hemorrhage. Chagot et al.(13) reported superficial wound infection in 3 out of 104 cases (2.9%), while Schuurman et al.(30). described two instances of symptomatic hardware infection requiring explantation. Isolated cases of lead malposition and perioperative seizure were also described (Hassan et al. and Oliveira et al.(21, 23)). Transient confusion, disequilibrium, or worsening gait were noted occasionally in early postoperative stages but generally resolved within weeks.

Two studies reporting surgical complication outcomes after DBS in MS were included in the analysis. (figure 3). The pooled complication rate was estimated at 11% (95% CI 0.00–1.00), although substantial heterogeneity was observed (I² = 91.0%, τ² = 3.6266, p = 0.0009). Individual study estimates were: Berk et al.(31): 4 events among 116 procedures (3%). Chagot et al.(13) 3 events among 12 procedures (33%). The large difference between studies contributed to the high heterogeneity observed in this analysis.

**Figure 3:**
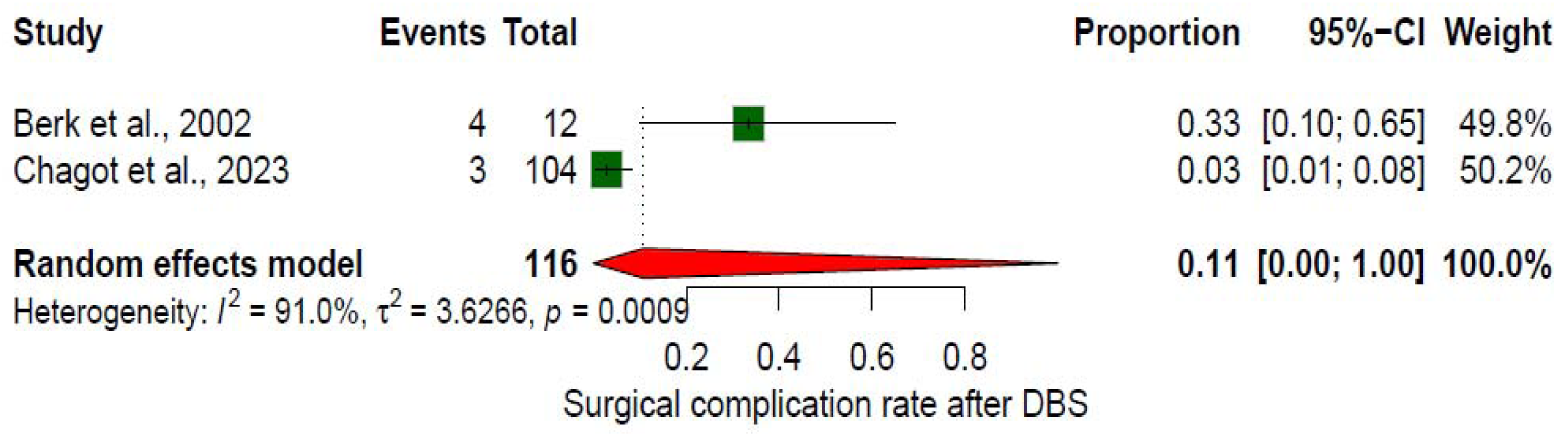
forest plot of surgical complication rate.

Device-related complications were observed in several cohorts during long-term follow-up. These included lead fracture or migration, connector displacement, pulse generator failure, and battery exhaustion necessitating replacement surgery. Hyam et al.(22) reported lead migration in one patient requiring revision; Banks et al. and Plaha et al.(5, 24), observed isolated hardware disconnections. Infection around the pulse generator site constituted the most frequent hardware issue, with pooled incidence of 5% (95% CI 2–10%), based on combined data from the infection forest plot. These events sometimes led to temporary discontinuation of stimulation or complete device removal. Two studies reported postoperative infection outcomes following DBS implantation. (figure 4). The pooled infection rate was estimated at approximately 7% (95% CI 0.00–1.00), with moderate to substantial heterogeneity (I² = 74.1%, τ² = 1.3468, p = 0.0496). Individual study estimates were: Berk et al.(31) 2 infections among 116 patients (approximately 2%). Chagot et al.(13): 3 infections among 12 patients (approximately 25%). The difference in infection rates between the two studies contributed to the observed heterogeneity.

**Figure 4:**
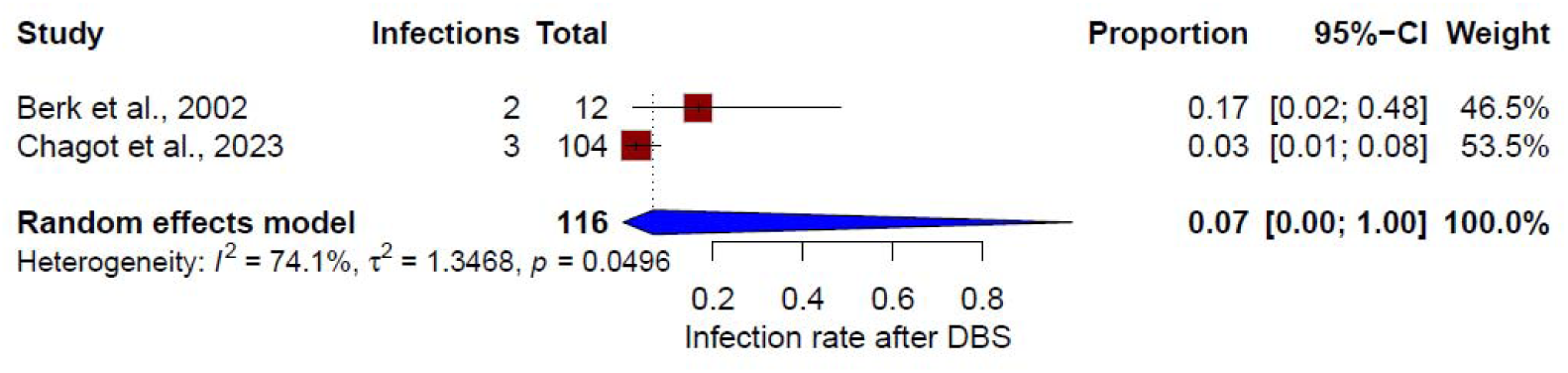
forest plot of infection rate after DBS.

Stimulation-induced side effects were far more common but typically reversible with parameter adjustment. Dysarthria, muscle contractions, paresthesia, dysesthesia, and disequilibrium were the most frequently reported transient side effects. Several studies (e.g., Hooper et al., Bittar et al. and Hassan et al.(21, 25, 28)) noted mild speech deterioration or imbalance during high-amplitude stimulation, which improved after lowering voltage or pulse width. Oliveira et al.(23) described variable tolerance among dual-lead thalamic implants, with off-target effects in cerebello-thalamic regions manifesting as worsening of ataxia. Plaha et al. and Hyam et al.(22, 24, 29) documented temporary dysmetria and limb heaviness during testing. Most side effects resolved promptly when stimulation parameters were modified. Table 3

### Risk of Bias and Quality Assessment

Risk of bias was assessed using the Joanna Briggs Institute (JBI) critical appraisal tools appropriate for each study design (Table 2). Across the included studies, methodological quality was generally moderate, with several domains insufficiently reported. The number of “Yes” ratings ranged from 5 to 8 across studies, indicating that most investigations fulfilled a moderate proportion of key methodological criteria. However, a considerable number of items were rated as “Unclear,” reflecting incomplete reporting of important methodological details. In particular, information regarding participant selection procedures, blinding of outcome assessors, and management of potential confounders was frequently insufficient or absent. Two studies were judged to have a higher overall risk of bias (Banks et al. Hassan et al.(5, 21)), primarily due to multiple “Unclear” ratings and limited methodological transparency. Most of the remaining studies were classified as having moderate risk of bias, while one cohort study (Hosseini et al.(26)) demonstrated comparatively stronger methodological quality with the highest number of “Yes” ratings and no domains judged as high risk. Although two randomized controlled trials (Oliveira et al. and Schuurman et al.(23, 29)) were included and showed acceptable methodological reporting, their multiple sclerosis subgroups were relatively small. The majority of other studies were observational or single-arm cohorts, which inherently limits internal validity and increases susceptibility to confounding. Overall, the risk-of-bias assessment indicates that the available evidence on deep brain stimulation for multiple sclerosis tremor is methodologically usable but constrained by several limitations, including small sample sizes, incomplete reporting of methodological safeguards, and limited control for confounding factors. Consequently, the pooled estimates derived from the meta-analysis should be interpreted with appropriate caution.

**Table 2:** Risk of Bias and Quality Assessment (JBI Critical Appraisal Tools)

| Study | Yes | No | Unclear | Not Applicable | Overall Risk |
| --- | --- | --- | --- | --- | --- |
| Banks et al., 2025 | 5 | 1 | 5 | 0 | High |
| Berk et al., 2002 | 6 | 0 | 5 | 0 | Moderate |
| Bittar et al., 2005 | 6 | 2 | 3 | 0 | Moderate |
| Chagot et al., 2023 | 6 | 1 | 4 | 0 | Moderate |
| Hassan et al., 2012 | 5 | 0 | 6 | 0 | High |
| Hooper et al., 2002 | 7 | 0 | 4 | 0 | Moderate |
| Hosseini et al., 2012 | 8 | 0 | 3 | 0 | Low |
| Hyam et al., 2007 | 7 | 1 | 3 | 0 | Moderate |
| Oliveria et al., 2017 | 7 | 0 | 4 | 0 | Moderate |
| Plaha et al., 2008 | 7 | 0 | 4 | 0 | Moderate |
| Schuurman et al., 2000 | 7 | 0 | 4 | 0 | Moderate |
| Schuurman et al., 2008 * | 7 | 0 | 4 | 0 | Moderate |

\* Note: The study by Schuurman et al., 2008 is a long-term follow-up of the original cohort from Schuurman et al., 2000. Therefore, its risk of bias scores reflects the methodological evaluation of the primary trial design.

**Table 3.**
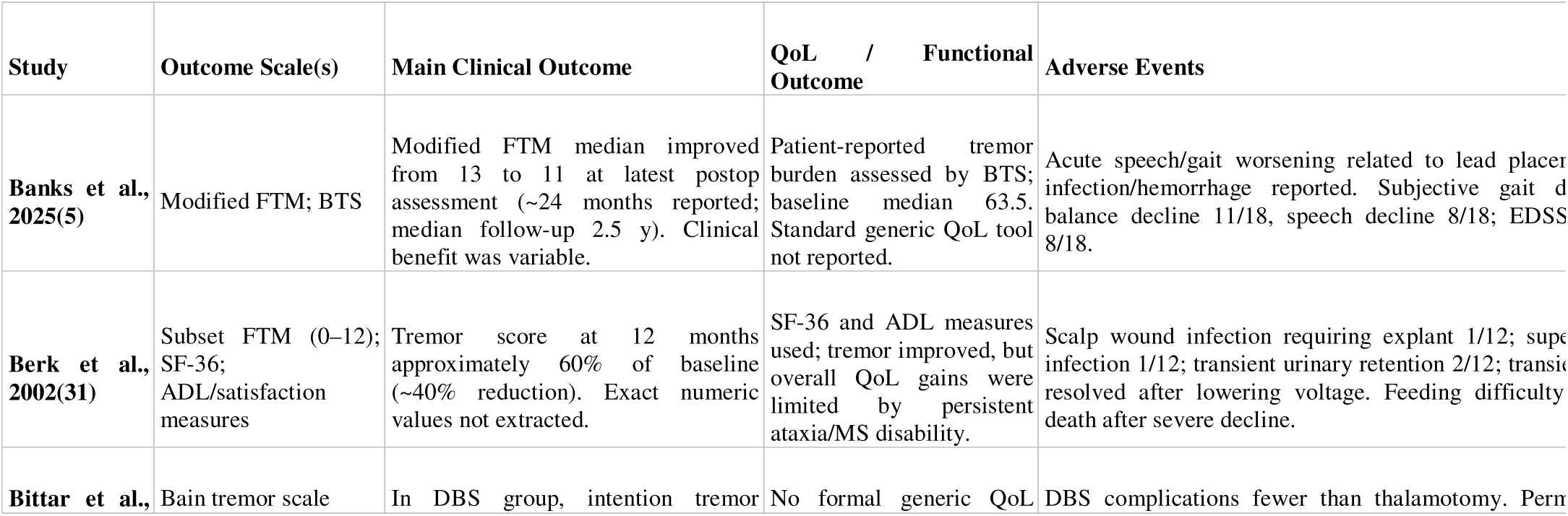
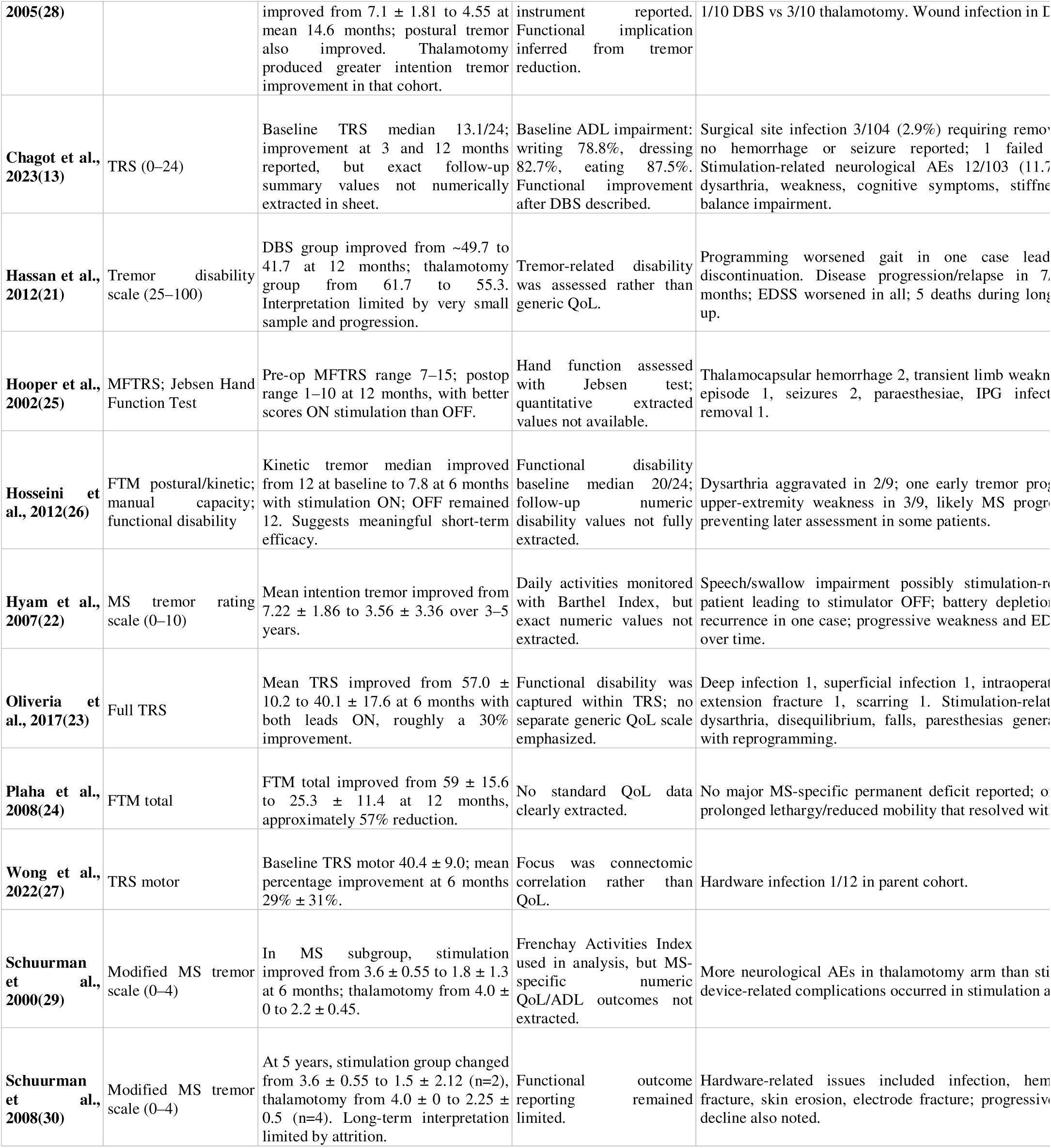
Clinical Outcomes and Adverse Events of DBS in MS Tremor.

### Publication bias

Potential publication bias was assessed using funnel plot visualization. The funnel plot demonstrated an approximately symmetrical distribution of effect sizes around the pooled estimate. Although the number of included studies was small, no obvious asymmetry suggestive of major publication bias was visually identified. (figure 5)

**Figure 5:**
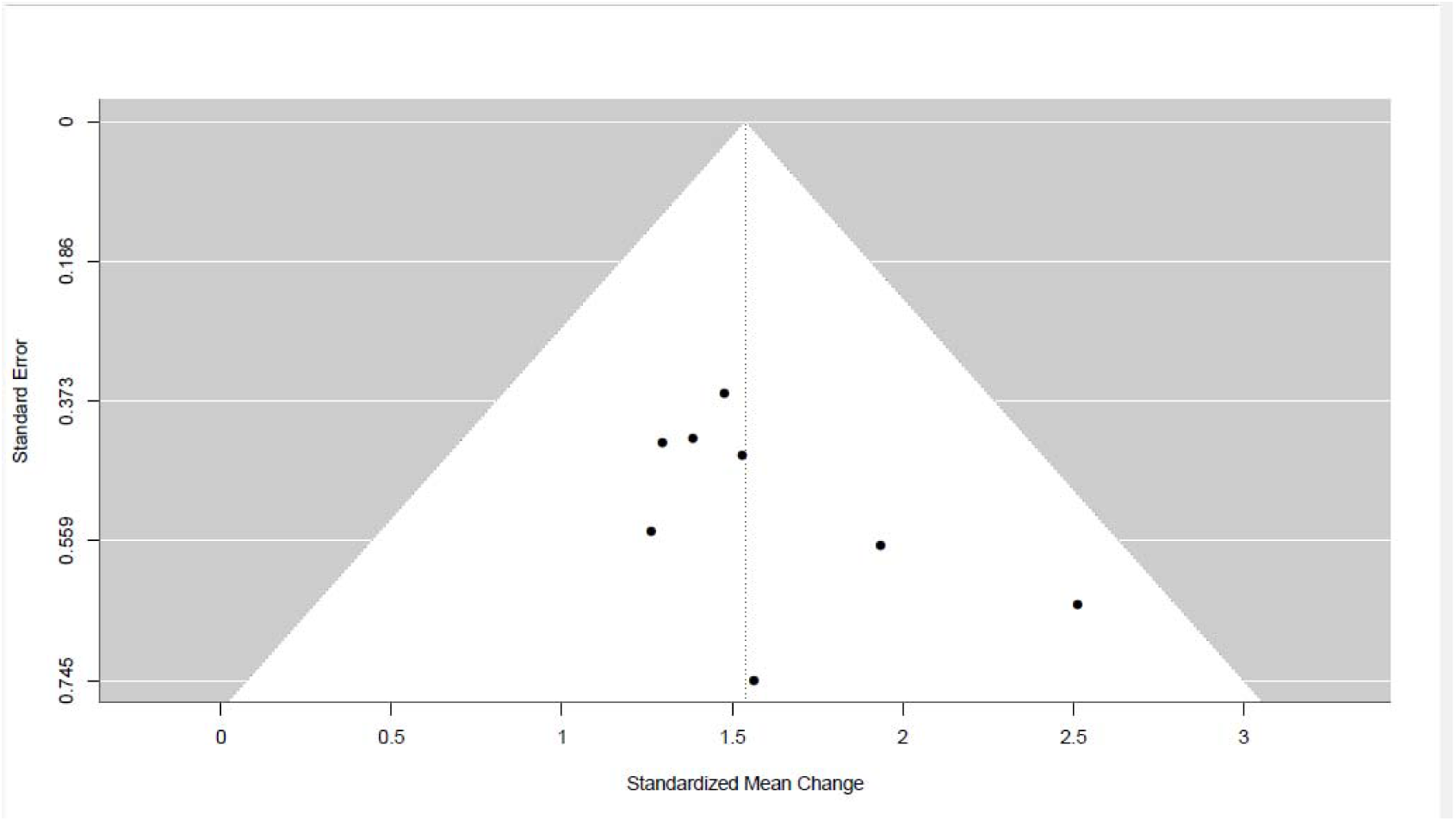
funnel plot of included studies in meta-analysis.

### Sensitivity analysis

Influence diagnostics and leave-one-out sensitivity analyses were performed to assess the robustness of the pooled effect estimate. No individual study demonstrated excessive influence on the pooled results across multiple diagnostic metrics, including studentized residuals, Cook’s distance, DFFITS, covariance ratios, and hat values. Removal of individual studies did not substantially alter the pooled estimate, indicating that the overall findings were not driven by a single influential study. (figure 6)

**Figure 6:**
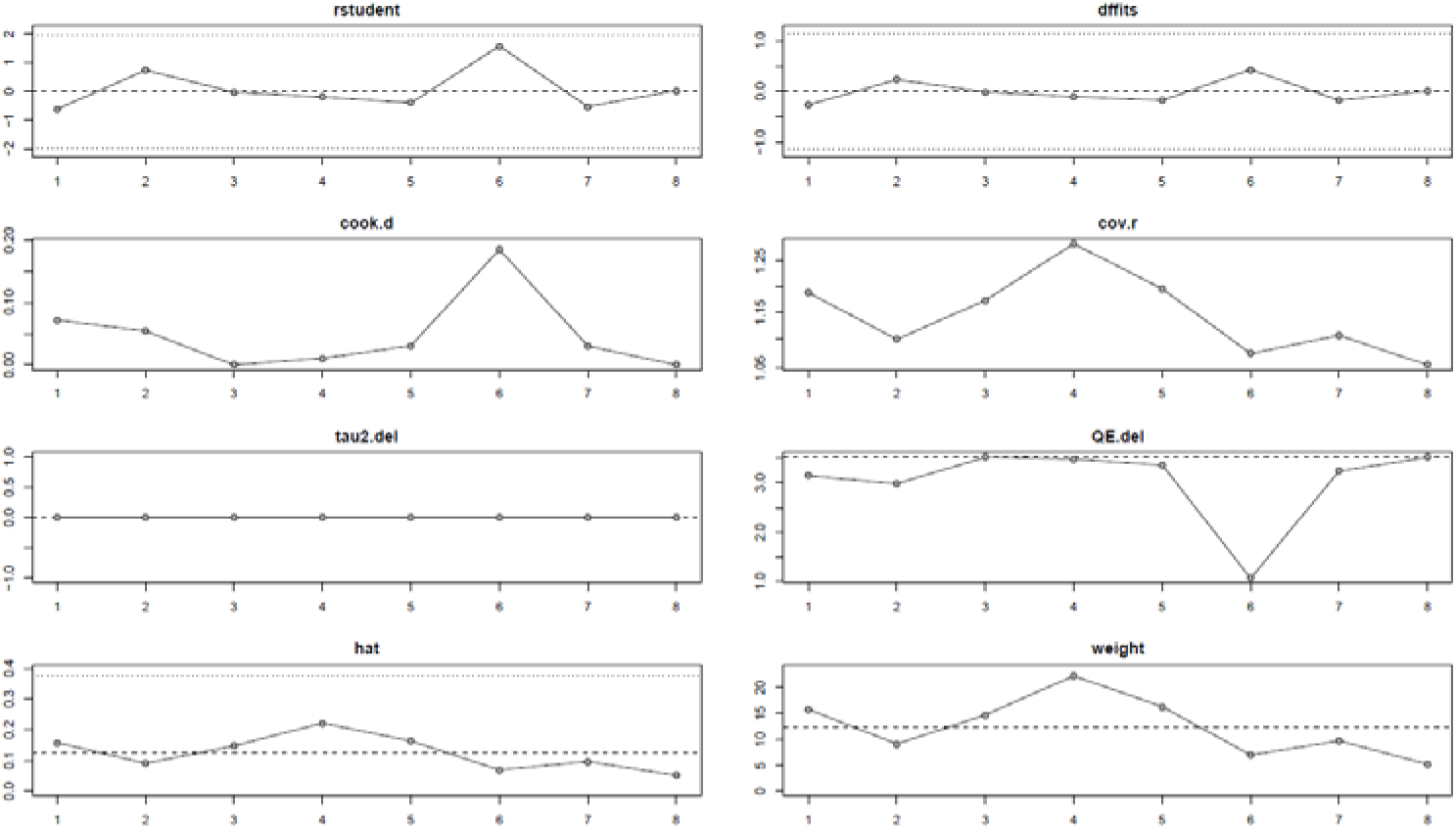
Leave-one sensitivity analysis.

### Meta-regression

An exploratory meta-regression analysis was performed to evaluate the relationship between study sample size and treatment effect. Visual inspection of the meta-regression plot suggested no clear association between sample size and effect size, indicating that the magnitude of tremor improvement following DBS was not systematically related to study size within the available dataset. (figure 7)

**Figure 7:**
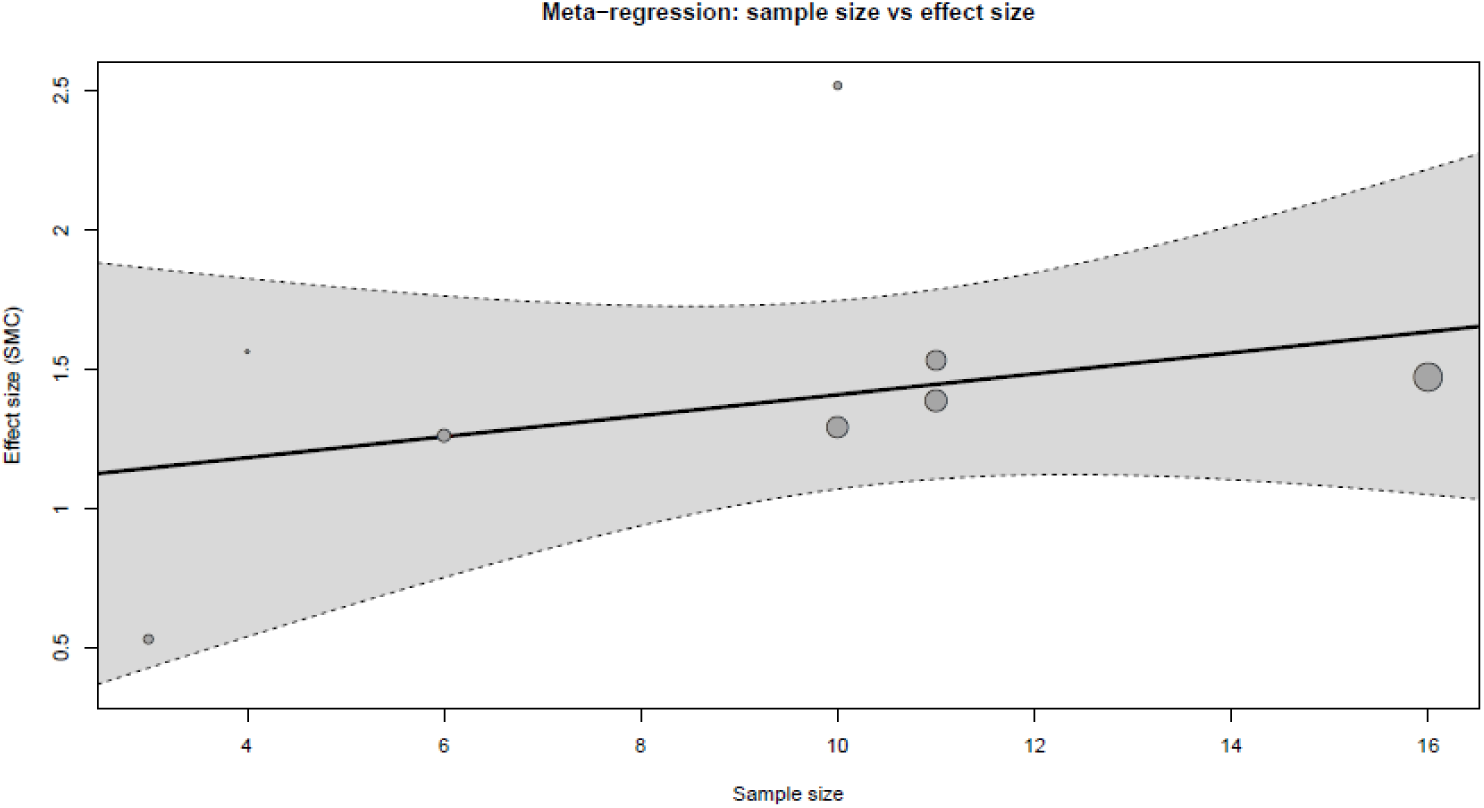
Exploratory Meta-Regression.

## Discussion

### Main findings

this systematic review and meta-analysis assessed deep brain stimulation (DBS) for tremor in people with multiple sclerosis (MS including 13 studies. Across the quantitative synthesis, eight studies contributed to the meta-analysis of tremor severity as primary clinical outcome. Overall, DBS was associated with a reduction in tremor severity, with a pooled standardized mean change (SMC) of 1.42 (95% CI 1.07 to 1.77) using a random-effects model with REML and Hartung–Knapp adjustment. From a heterogeneity perspective, the pooled analysis showed I² = 0.0% (τ² < 0.0001; p=0.6300), indicating no observed between-study statistical heterogeneity. However, this should be interpreted cautiously: the number of available studies was limited, and multiple methodological differences across included cohorts (e.g., DBS targets including predominantly VIM with occasional alternative/combined targets; unilateral versus bilateral implantation; and follow-up duration ranging from months to over two decades) may limit the ability of the analysis to detect true variability in treatment effects. Consistent direction of benefit was observed across individual studies, with effect sizes ranging from 1.26 to 2.51. Secondary outcomes are adverse and safety-related events. Device-related complications were reported during longer-term follow-up, including lead migration or fracture, connector displacement, and pulse generator-related failure requiring revision or replacement surgery; infection around the pulse generator site was described as an important event type. In the infection analysis based on two studies, the pooled infection rate was approximately 7% (95% CI 0.00–1.00), with substantial heterogeneity (I² = 74.1%, τ² = 1.3468; p=0.0496), driven by discordant infection rates between studies (e.g., 2/116 vs 3/12 infections). In addition to infectious complications, stimulation-induced adverse effects such as dysarthria, disequilibrium and other transient side effects were reported and were generally reversible with parameter adjustments; ataxia-related worsening (off-target effects) was also described in some cohorts. Finally, risk-of-bias assessment using Joanna Briggs Institute tools suggested evidence of methodological limitations across studies (frequent “Unclear” domains related to participant selection, assessor blinding, and confounder management, and two studies judged higher risk), which further supports cautious interpretation of pooled estimates.

### Comparison with Previous Studies

The findings of the present study are generally compared with previous studies evaluating the role of deep brain stimulation (DBS) in the management of multiple sclerosis (MS)–related tremor, although important methodological differences exist. The most comprehensive prior synthesis was conducted by Zali et⍰al.(12), who included 17 studies published between 1999 and 2018 with 168 patients. Their analysis reported a pooled tremor improvement rate of 73% (95% CI 64–83%) and a standardized mean difference (SMD) of −2.9, suggesting a substantial reduction in tremor severity following DBS. These results support the overall therapeutic potential of DBS for medically refractory MS tremor and align with the general direction of our findings. However, a key methodological limitation of the Zali et⍰al. meta-analysis was the substantial statistical heterogeneity reported in their pooled analyses (I²⍰=⍰84.1% for tremor improvement and I²⍰=⍰89.8% for SMD). As highlighted in a subsequent methodological correspondence by Serhan et⍰al.(32), several sources of variability may explain this heterogeneity. The included studies differed markedly in sample sizes, with some trials enrolling fewer than five participants while others included more than fifteen patients, which may inflate effect size estimates and reduce reproducibility. In addition, there was considerable variability in DBS parameters, including stimulation targets, electrode configuration, stimulation frequency and amplitude, and duration of follow-up. Such methodological diversity may limit the clinical interpretability of pooled estimates and underscores the need for more standardized study designs. Another methodological issue raised in the same correspondence relates to the use of the Cochrane risk-of-bias tool in the Zali et⍰al. review. Because most of the included studies were single-arm trials or case-series–like designs rather than randomized controlled trials, the Cochrane tool may not have been the most appropriate instrument for quality assessment. In contrast, our study applied the JBI Critical Appraisal Tools, which are better suited for evaluating observational and case-series evidence. Our risk-of-bias assessment demonstrated that most included studies were of moderate quality, with one study classified as low risk of bias (Hosseini et⍰al.(26)), two studies categorized as high risk (Banks et⍰al. and Hassan et⍰al.(5, 21)), and the remainder showing moderate methodological quality. This structured assessment provides a clearer evaluation of the evidence base and highlights the ongoing limitations in study design within this field.

The present findings should also be interpreted within the broader neuromodulation literature in multiple sclerosis. An umbrella review by Yaseri et⍰al.(33), which synthesized evidence from 14 systematic reviews of both invasive and non-invasive brain stimulation techniques, concluded that DBS appears effective in reducing tremor in patients with MS, although the overall evidence remains limited and heterogeneous. The authors emphasized that differences in stimulation techniques, treatment parameters, and outcome measures remain major limitations to conclude definitive conclusions, emphasizing the need for standardized protocols in future research. Evidence from other movement disorder populations also provides useful context for interpreting DBS outcomes. For example, a recent network and multilevel meta-analysis(34) comparing ventral intermediate nucleus (VIM) and posterior subthalamic area (PSA) stimulation for essential tremor included 52 studies and demonstrated that both targets produce substantial tremor reduction, with mean differences of −28.24 and −31.23 in tremor rating scores, respectively, without a statistically significant difference between targets (p⍰=⍰0.428). in mentioned review, PSA stimulation was associated with greater improvement in head tremor and a lower incidence of dysarthria, which may be due to anatomical proximity of cerebellothalamic pathways such as the dentato-rubro-thalamic tract. These findings are relevant to MS tremor because similar cerebello-thalamo-cortical circuits are believed to contribute to tremor generation in MS.

Overall, the existing literature consistently suggests that DBS can provide clinically meaningful tremor reduction in patients with MS. Nevertheless, previous studies have been limited by small sample sizes, heterogeneous methodologies, and inconsistent reporting of stimulation parameters and outcomes. By incorporating updated evidence, applying an appropriate risk-of-bias assessment framework, and contextualizing our findings within the broader neuromodulation literature, the present study contributes to a more rigorous synthesis of the available evidence and highlights the need for larger, standardized clinical investigations in this field.

### Target Selection and Neuroanatomical Considerations

Selection of the optimal stimulation target is a critical determinant of clinical outcomes in deep brain stimulation (DBS) for tremor disorders. In patients with multiple sclerosis (MS), tremor is thought to arise from disruption of the cerebello–thalamo–cortical circuitry due to demyelinating lesions affecting the cerebellum and its output pathways (Alusi et⍰al.; Bayoumi et⍰al.; Zhang et⍰al.(6, 7, 35)). Structural and diffusion-based studies have demonstrated that lesions involving the cerebellar white-matter tracts and the dentato-rubro-thalamic pathway can produce abnormal activity within thalamocortical networks, which is believed to underlie the generation of intention and postural tremor commonly observed in MS (Bayoumi et⍰al.; Zhang et⍰al.(6, 35)). Because of this network-level pathophysiology, neuromodulation targeting key relay nodes within the cerebellothalamic system has become the principal surgical strategy for medically refractory MS tremor. Historically, the ventral intermediate nucleus (VIM) of the thalamus has been the most commonly targeted structure for DBS in tremor syndromes, including MS-associated tremor (Zali et⍰al.; Kalogirou et⍰al.(10, 12)). The VIM receives afferent projections from the contralateral cerebellar dentate nucleus and functions as a major relay within the cerebello–thalamo–cortical network, transmitting cerebellar output to motor cortical regions involved in voluntary movement control. (9, 11, 29, 34). Clinical studies have shown that VIM stimulation can significantly reduce tremor severity and improve functional outcomes in patients with severe, medication-refractory MS tremor (Berk et⍰al.; Hooper et⍰al.; Hosseini et⍰al.; Zali et⍰al.(12, 25, 26, 31)). However, therapeutic responses remain heterogeneous, likely reflecting the complex and multifocal nature of MS-related network disruption. In recent years, increasing attention has been directed toward alternative targets located along the cerebellothalamic pathway. The posterior subthalamic area (PSA), including the zona incerta (ZI) and the prelemniscal radiation, contains fibers of the dentato-rubro-thalamic tract (DRTT), a major conduit of cerebellar output involved in tremor propagation (Zhang et⍰al.(35)). Stimulation of the ZI has been proposed to exert broader network-modulating effects through GABAergic projections that influence both basal ganglia–thalamo-cortical and cerebellar motor circuits (Zhang et⍰al.(35)). Because of these extensive connections, neuromodulation in this region may suppress tremor by modulating distributed oscillatory networks rather than a single thalamic relay nucleus. Evidence from comparative neuromodulation literature further supports the relevance of these alternative targets. A recent network and multilevel meta-analysis including 52 studies demonstrated that both VIM and PSA DBS provide substantial tremor reduction with comparable overall efficacy (mean tremor score reduction −28.24 vs −31.23; p⍰=⍰0.428), although PSA stimulation was associated with greater improvement in head tremor and a lower risk of dysarthria (Aghajanian et⍰al.(34)). Importantly, this analysis also demonstrated a gradual decline in DBS efficacy over time, estimated at approximately 1.2 tremor score points per year, highlighting the progressive nature of tremor disorders and the need for long-term follow-up (Aghajanian et⍰al.(34)). These findings suggest that stimulation closer to cerebellothalamic fiber pathways may influence tremor circuits more selectively while potentially reducing certain stimulation-related adverse effects. Another important consideration is the laterality of stimulation. Evidence from movement disorder literature indicates that unilateral DBS can produce substantial improvement in contralateral tremor symptoms, whereas bilateral implantation may provide broader control of axial or midline tremors but at the cost of increased adverse effects such as dysarthria and gait disturbance (Vetkas et⍰al.(36)). Therefore, individualized treatment strategies that consider symptom distribution, functional disability, and patient characteristics are essential. The heterogeneity of tremor mechanisms in MS further complicates target selection. Demyelinating lesions may affect multiple nodes within the cerebello–thalamo–cortical network, and concomitant cerebellar ataxia or dysmetria may limit the functional benefits achieved through tremor suppression alone (Alusi et⍰al.; Zhang et⍰al.(7, 35)). Consequently, careful patient selection remains a critical determinant of surgical success. Previous studies have emphasized that patients with disabling upper-limb tremor, relatively stable disease status, and limited cerebellar ataxia are more likely to benefit from DBS interventions (Paranathala (14)et⍰al.; Zali et⍰al.(12)).

Overall, current evidence suggests that although VIM stimulation remains the traditional and most widely applied target for MS-related tremor, alternative targets such as the PSA or ZI may provide comparable tremor suppression with potentially distinct side-effect profiles. Advances in connectomic imaging, tractography-guided targeting, and network-based neuromodulation approaches may help refine patient-specific targeting strategies and improve long-term outcomes in this complex and heterogeneous patient population.(9, 11, 34).

### Safety and adverse events

Safety outcomes following deep brain stimulation for multiple sclerosis–related tremor were inconsistently reported across the available literature. Although most studies primarily focused on tremor reduction, information regarding procedure-related complications and device-related adverse events was frequently limited or incompletely described. In the present study, the pooled infection rate was approximately 7%, with considerable heterogeneity between studies. This variability likely reflects differences in surgical techniques, perioperative management protocols, follow-up duration, and reporting practices across centers. The reported complications in the literature include wound infection, hardware-related complications, stimulation-induced dysarthria, gait instability, paresthesia, and transient disequilibrium. Long-term stimulation-related side effects such as speech disturbance and gait impairment have been widely reported in DBS literature for tremor disorders, particularly in thalamic stimulation targeting the ventral intermediate nucleus (VIM). These effects are often related to current spread to adjacent cerebellothalamic or corticobulbar pathways. Interpretation of adverse events in patients with multiple sclerosis requires particular caution. Many patients already present with baseline cerebellar dysfunction, dysarthria, or balance impairment due to the underlying disease process. Consequently, distinguishing between stimulation-related adverse effects and manifestations of disease progression can be challenging. Previous studies have emphasized that while DBS can significantly improve tremor severity, it generally does not improve cerebellar ataxia, which may limit functional benefit in some patients.

Overall, the current evidence suggests that DBS is relatively safe in carefully selected patients with severe medication-refractory MS tremor. However, the limited number of studies reporting safety outcomes and the lack of standardized complication reporting frameworks highlight the need for more systematic documentation of adverse events in future clinical investigations.

### Limitations

Several limitations should be considered when interpreting the findings of this meta-analysis.

First, the number of available studies evaluating deep brain stimulation for tremor in patients with multiple sclerosis remains relatively small. Only a limited number of studies met the eligibility criteria for quantitative synthesis of tremor outcomes, and an even smaller number provided detailed information on safety outcomes such as postoperative infections or device-related complications. The small evidence base restricts the statistical power of additional analyses such as subgroup comparisons or meta-regression and limits the robustness of publication bias assessment.

Second, the majority of included studies were observational cohort studies without randomized control groups. The absence of randomized controlled trials increases susceptibility to selection bias and confounding factors and limits the ability to establish definitive causal relationships between DBS treatment and clinical outcomes. This limitation reflects the broader challenge in the field of neurosurgical interventions, where randomized trials are often difficult to conduct.

Third, sample sizes in many of the included studies were relatively small. Several studies included fewer than ten patients and in some reports the sample size was five or fewer participants. Small cohorts increase the risk of imprecise effect estimates and may inflate reported treatment effects. In addition, small sample sizes may amplify study-level variability and reduce the generalizability of findings to the broader population of patients with MS tremor. Furthermore, although statistical heterogeneity in the primary meta-analysis was low, the small number of included studies may have limited the statistical power to detect true between-study variability.

Fourth, substantial clinical heterogeneity exists across studies in terms of patient characteristics and disease profiles. Patients differed in MS subtype, disease duration, baseline disability, tremor distribution, and the presence of cerebellar signs such as ataxia or dysmetria. Because tremor in multiple sclerosis arises from complex and heterogeneous disruptions of cerebello-thalamo-cortical circuits, these differences in patient characteristics may influence responsiveness to DBS and contribute to variability in treatment outcomes.

Fifth, there was variability in surgical targets and stimulation parameters among the included studies. Although most investigations targeted the ventral intermediate nucleus of the thalamus, other studies explored adjacent cerebellothalamic pathways or alternative stimulation strategies. Differences in electrode placement, stimulation amplitude, pulse width, frequency, and programming strategies may influence both efficacy and adverse-event profiles, thereby introducing additional heterogeneity across studies.

Sixth, outcome measurement was not fully standardized across studies. Tremor severity was assessed using different rating instruments, including variations of the Fahn–Tolosa–Marin tremor rating scale and other clinical tremor assessment tools. Although statistical standardization methods allow pooling across different scales, differences in measurement instruments and scoring systems may still introduce methodological variability.

Seventh, follow-up duration varied substantially across the included studies. Some studies evaluated outcomes within months after surgery, whereas others reported longer follow-up periods extending several years. Because multiple sclerosis is a progressive neurodegenerative disease, long-term outcomes following DBS may differ from early postoperative improvements. Variability in follow-up intervals therefore complicates direct comparison between studies.

Eighth, reporting of adverse events and complications was inconsistent across studies. Several studies primarily focused on tremor outcomes and provided only limited information on surgical complications or stimulation-related adverse effects. This incomplete reporting restricts the ability to draw firm conclusions regarding the safety profile of DBS in this patient population.

Finally, the relatively small number of included studies limits the reliability of funnel plot–based assessments of publication bias. When fewer than ten studies are available, statistical tests for publication bias have limited sensitivity and should therefore be interpreted with caution.

Taken together, these limitations highlight the need for larger prospective studies with standardized outcome measures, clearly defined follow-up intervals, and comprehensive reporting of both efficacy and safety outcomes in patients undergoing DBS for multiple sclerosis–related tremor.

### Clinical implications and future directions

Despite the limitations of the current evidence base, several clinically relevant conclusions can be drawn. Deep brain stimulation appears to provide meaningful tremor reduction in selected patients with severe, medication-refractory MS tremor. Given the substantial impact of tremor on activities of daily living such as writing, eating, and self-care, successful tremor control may translate into meaningful improvements in functional independence.

However, careful patient selection remains essential. Tremor in multiple sclerosis often occurs in the context of cerebellar dysfunction, and patients with severe ataxia or dysmetria may experience limited functional improvement despite reduction in tremor amplitude.

Comprehensive preoperative evaluation including neurological examination and functional assessment is therefore critical when considering surgical intervention.

### Conclusion

This systematic review and meta-analysis suggests that deep brain stimulation (DBS) may reduce tremor severity in patients with multiple sclerosis. Across eight studies included in the quantitative synthesis, DBS was associated with an overall improvement in tremor scores (SMC⍰=⍰1.42; 95%⍰CI⍰1.07–1.77). Device-related complications and stimulation-induced side effects were reported, including hardware problems and infections, with a pooled infection rate of approximately 7%, while most stimulation-related symptoms were transient and responsive to programming adjustments. However, the current evidence base remains limited. The included studies were small and methodologically heterogeneous, with variability in stimulation targets, implantation strategies, follow-up duration, and outcome reporting. In addition, the absence of observed statistical heterogeneity (I²⍰=⍰0%) should be interpreted cautiously given the limited number of studies. Larger prospective studies with standardized outcome measures and more consistent reporting of safety outcomes are needed to better define the long-term role of DBS in the management of MS-related tremor.

## Declaration of Interest

The authors declare no competing interests.

## Declaration of AI Use

Artificial intelligence (AI) tools (ChatGPT) were used to improve the grammar, clarity, and language of this manuscript. The authors reviewed and approved all content, and take full responsibility for the integrity and accuracy of the work.

## Supporting information

extraction

rob

prisma checklist

protocol

full text screening 1

full text screening 2

search strategy

title abstract screening

prisma flowchart

## Data Availability

All data produced in the present study are available upon reasonable request to the authors

## Acknowledgments

The authors have no acknowledgments to declare.

## Funding

This research received no external funding.

## Appendix and Supplementary Material

- Appendix 1. PRISMA checklist
- Appendix 2. Prospero Protocol
- Appendix 3. Search Strategy
- Appendix 4. Title-Abstract screening excluded studies
- Appendix 5. Full text screening Excluded and Included studies
- Appendix 6. Data extraction Sheets
- Appendix 7. Risk of bias

## Ethics Approval and Consent to Participate

This study is a systematic review and meta-analysis based exclusively on previously published studies. No primary data were collected and no direct involvement of human participants occurred. Therefore, ethical approval from an Institutional Review Board (IRB) or Ethics Committee was not required.

**Consent to Participate declaration in the manuscript**: Not applicable.

**Human Ethics and Consent to Participate declarations:** Not applicable.

**Clinical trial number:** not applicable.

## Notes

### Competing Interest Statement

The authors have declared no competing interest.

### Funding Statement

This study did not receive any funding

