## Supplementary material for "Therapeutic Efficacy and Safety of Deep Brain Stimulation for Multiple Sclerosis Related-Tremor: A Systematic Review and Meta-Analysis": protocol

NIHR | National Institute for Health and Care Research

PROSPERO
International prospective register of systematic reviews

### Therapeutic efficacy and safety of deep brain stimulation for multiple sclerosis–related tremor: a systematic review and meta-analysis

*Farzan Fahim, Amirmahdi Mojtahedzadeh*

**Citation**
Farzan Fahim, Amirmahdi Mojtahedzadeh. Therapeutic efficacy and safety of deep brain stimulation for multiple sclerosis–related tremor: a systematic review and meta-analysis. PROSPERO 2026 CRD420261347426. Available from https://www.crd.york.ac.uk/PROSPERO/view/CRD420261347426.

#### REVIEW TITLE AND BASIC DETAILS

##### Review title
Therapeutic efficacy and safety of deep brain stimulation for multiple sclerosis–related tremor: a systematic review and meta-analysis

##### Condition or domain being studied
*Deep Brain Stimulation; Multiple Sclerosis; Tremor*

##### Rationale for the review
Multiple sclerosis is a chronic disabling disease associated with sensory and visual symptoms; however, tremor is one of the most disabling manifestations and can markedly interfere with patients' daily activities. Medical management often fails to adequately control MS-related tremor. With advances in understanding the pathophysiological neural networks involved in tremor generation, neuromodulatory interventions such as deep brain stimulation (DBS) have been introduced. Therefore, we aimed to comprehensively review all original studies to assess the therapeutic effect of DBS in reducing tremor in patients with multiple sclerosis, as well as to evaluate the safety of this procedure.

##### Review objectives
1. what is the impact of deep brain stimulation to control tremor related MS?

2. is deep brain stimulation is safe in patients with tremor related MS?

##### Keywords
Deep Brain Stimulation; Multiple sclerosis; Essential tremor; Systematic review

##### Country
Iran (Islamic Republic of)

#### ELIGIBILITY CRITERIA

##### Population

*Included*
patients who suffering from tremor-related MS

##### Intervention(s) or exposure(s)

*Included*
mentioned patients who undergo deep brain stimulation.

##### Comparator(s) or control(s)

*Included*
*PICO tags selected: Medical Therapy; Sham Intervention ; Thalamotomy*

##### Study design
Both randomized and nonrandomized study types will be included.

*Included*
clinical trials

cohort studies

##### Context
This review focuses on clinical studies evaluating deep brain stimulation (DBS) for the treatment of tremor associated with multiple sclerosis (MS). Eligible studies include original clinical investigations conducted in neurosurgical centers where DBS was performed for patients with medically refractory MS-related tremor. Participants were adults diagnosed with multiple sclerosis who experienced disabling tremor that significantly affected daily activities and had insufficient response to medical therapy.

The intervention of interest is deep brain stimulation targeting tremor-related neural circuits. Studies were included if they reported quantitative tremor outcomes before and after DBS implantation, allowing calculation or direct extraction of standardized mean change (SMC) in tremor severity. Both studies reporting direct statistical measures and studies requiring data conversion were considered eligible.

The review synthesizes evidence from available original studies that measured tremor outcomes following DBS implantation and reported sufficient data for quantitative meta-analysis. These studies were conducted in clinical neurosurgical settings and evaluated the therapeutic effectiveness of DBS for MS-related tremor, as well as procedure-related safety outcomes such as complications and infections.

#### TIMELINE OF THE REVIEW

##### Date of first submission to PROSPERO
21 March 2026

##### Review timeline
Start date: 21 March 2026. End date: 21 June 2026.

##### Date of registration in PROSPERO
21 March 2026

#### AVAILABILITY OF FULL PROTOCOL

##### Availability of full protocol
A full protocol has not been written.

#### SEARCHING AND SCREENING

##### Search for unpublished studies
Both published and unpublished studies will be sought.

##### Main bibliographic databases that will be searched
The main databases to be searched are *MEDLINE*, *PubMed* and *Scopus*.

*Other important or specialist databases that will be searched*
embase ( not via ovid)

web of science

##### Search language restrictions
There are no language restrictions.

##### Search date restrictions
There are no search date restrictions.

##### Other methods of identifying studies
Other studies will be identified by: *reference list checking (backward citation searching)* and *searching trial or study registers*.

##### Link to search strategy
A full search strategy has been uploaded to PROSPERO. The PDF may be accessed through this link https://www.crd.york.ac.uk/PROSPEROFILES/e72b3111dd873f99b7410743840fcbba.pdf.

##### Selection process

Studies will be screened independently by at least two people (or person/machine combination) with a process to resolve differences.

##### Other relevant information about searching and screening

None

#### DATA COLLECTION PROCESS

##### Data extraction from published articles and reports

Data will be extracted independently by at least two people (or person/machine combination) with a process to resolve differences.

Authors will not be contacted for further information.

##### Study risk of bias or quality assessment

Risk of bias will be assessed using:

jbi risk assessment checklists

Data will be assessed independently by at least two people (or person/machine combination) with a process to resolve differences.

Additional information will be sought from study investigators if required information is unclear or unavailable in the study publications/reports.

##### Reporting bias assessment

Risk of bias due to missing results will be assessed

##### Certainty assessment

Certainty of findings will not be assessed

#### OUTCOMES TO BE ANALYSED

##### Main outcomes

main outcome is tremor controlling by dbs

##### Additional outcomes

1. complication of procedure

2. infection rate

#### PLANNED DATA SYNTHESIS

##### Strategy for data synthesis

Quantitative data synthesis will be conducted using meta-analysis of effect sizes extracted from the included primary studies. Because most eligible studies evaluating deep brain stimulation (DBS) for multiple sclerosis–related tremor report outcomes using pre- and post-intervention tremor scores, the primary effect measure will be the standardized mean change (SMC), which reflects the magnitude of tremor reduction after DBS implantation.

For each study, effect sizes and corresponding 95% confidence intervals will be extracted directly from the reported results or calculated from available summary statistics when necessary. When studies do not directly report SMC values, the effect size will be derived from the reported pre- and post-treatment means and standard deviations using established methods for standardized mean change estimation.

Pooled effect estimates will be calculated using a random-effects meta-analysis model with the Hartung–Knapp adjustment and restricted maximum likelihood (REML) estimator to account for potential clinical and methodological variability among studies. The overall pooled SMC will represent the magnitude of tremor improvement following DBS across all included studies.

Statistical heterogeneity will be assessed using the $I^2$ statistic, $\tau^2$, and the Cochran Q test. Based on preliminary pooled analysis of the included studies, heterogeneity is expected to be low ($I^2 \approx 0\%$), suggesting consistent treatment effects across studies.

Subgroup analyses will be conducted according to the method of effect size derivation. Specifically, studies will be categorized into two groups: (1) studies in which effect sizes are directly reported or directly calculable from published statistics, and (2) studies in which the effect sizes require conversion or indirect estimation from available data. Separate pooled estimates will be calculated for each subgroup, and statistical differences between subgroups will be evaluated using a chi-square test for subgroup differences.

Forest plots will be generated to visually present the pooled estimates and individual study effects. Additional analyses may include sensitivity analyses to evaluate the robustness of the pooled results. All statistical analyses will be performed using appropriate meta-analysis software and standard methodological approaches for quantitative evidence synthesis.

#### CURRENT REVIEW STAGE

##### Stage of the review at this submission

| Review stage | Started | Completed |
|---|---|---|
| Pilot work | ✓ | ✓ |
| Formal searching/study identification | ✓ | ✓ |
| Screening search results against inclusion criteria | ✓ | ✓ |
| Data extraction or receipt of IPD | | |
| Risk of bias/quality assessment | | |

| Review stage | Started | Completed |
| --- | --- | --- |
| Data synthesis | | |

##### Review status

The review is currently planned or ongoing.

##### Publication of review results

Results of the review will be published.

#### REVIEW AFFILIATION, FUNDING AND PEER REVIEW

##### Review team members

**Dr Farzan Fahim** (review guarantor and contact) ORCID: 0000-0003-0591-674X. Shohada-E-Tajrish Hospital. Iran.

No conflict of interest declared.

**Dr Amirmahdi Mojtahedzadeh**. Shahid Beheshti University of Medical Science. Iran.

No conflict of interest declared.

##### Named contact

##### Review affiliation

shahid beheshti university of medical science

##### Funding source

Review has no funding and no agreed support from an academic institution and is done in authors' own time.

##### Peer review

There has been no peer review of this planned review.

#### ADDITIONAL INFORMATION

##### Additional information

This review aims to provide a quantitative synthesis of the available clinical evidence on the effectiveness of deep brain stimulation (DBS) for tremor associated with multiple sclerosis. To our knowledge, previous literature on this topic has largely consisted of narrative reviews or small case series, and few studies have quantitatively synthesized the magnitude of tremor improvement across available clinical studies. The present review therefore focuses on systematically extracting pre- and post-intervention tremor outcomes from published clinical studies and estimating standardized mean change (SMC) as the primary effect measure.

A distinguishing methodological aspect of this review is the use of standardized mean change derived from pre- and post-treatment tremor scores to quantify the magnitude of therapeutic improvement following DBS implantation. Studies reporting sufficient summary statistics for direct calculation of effect size will be analyzed separately from studies in which effect sizes must be derived through statistical conversion from reported data. Accordingly, subgroup analyses will be conducted based on the method of effect size derivation (direct extraction versus converted estimates) in order to evaluate whether the data extraction approach influences the pooled treatment effect.

This review is not commissioned by any external organization or policy-making body. The study is conducted as an independent academic research project aimed at improving the quantitative understanding of DBS effectiveness for tremor in patients with multiple sclerosis. No additional partner organizations are formally involved in the conduct of the review beyond the listed review team members.

##### Review conflict of interest

Declared individual interests are recorded under team member details.. No additional interests are recorded for this review.

##### Medical Subject Headings

Deep Brain Stimulation; Multiple Sclerosis

#### SIMILAR REVIEWS

##### Check for similar records already in PROSPERO

*PROSPERO identified a number of existing PROSPERO records that were similar to this one (last check made on 21 March 2026). These are shown below along with the reasons given by that the review team for the reviews being different and/or proceeding.*

- Efficacy of unilateral and bilateral ventral intermediate nucleus deep brain stimulation for essential tremor: a systematic review and meta-analysis [published 23 March 2020] [CRD42020140830]. The review was judged **not to be similar**
- Systematic review and meta-analysis of voice outcomes following Thalamus -deep brain stimulation in tremor. [published 15 October 2023] [CRD42023462225]. The review was judged **not to be similar**
- Systematic Review of Deep Brain Stimulation Targets and Stimulation Methods in Essential Tremor [published 2 October 2023] [CRD42023466284]. The review was judged **not to be similar**
- The outcomes and complications of deep brain stimulation in the treatment of essential tremor: a single group meta-analysis [published 4 February 2020] [CRD42020147313]. The review was judged **not to be similar**
- Non-motor symptoms in subthalamic nucleus deep brain stimulation: a meta-analysis [published 18 September 2019] [CRD42019133932]. The review was judged **not to be similar**

#### PROSPERO version history

- Version 1.0, published 21 Mar 2026

#### Disclaimer

The content of this record displays the information provided by the review team. PROSPERO does not peer review registration records or endorse their content.

PROSPERO accepts and posts the information provided in good faith; responsibility for record content rests with the review team. The guarantor for this record has affirmed that the information provided is truthful and that they understand that deliberate provision of inaccurate information may be construed as scientific misconduct.

PROSPERO does not accept any liability for the content provided in this record or for its use. Readers use the information provided in this record at their own risk.

Any enquiries about the record should be referred to the named review contact
