## Supplementary material for "Therapeutic Efficacy and Safety of Deep Brain Stimulation for Multiple Sclerosis Related-Tremor: A Systematic Review and Meta-Analysis": search strategy

Supplementory of Search Strategy

Pubmed :188

No limitation of time and language

("multiple sclerosis" OR "MS" OR "demyelinatingdisease*" OR "demyelination disorder*") AND ("deep brainstimulation" OR DBS OR "neuromodulation" OR "brainstimulation" OR "stereotactic stimulation" OR "implantableneurostimulator" OR "thalamic stimulation" OR "VIMstimulation" OR "ventral intermediate nucleus stimulation") AND(tremor OR "intention tremor" OR "postural tremor" OR"action tremor" OR "MS tremor" OR "cerebellartremor")

________________________________________________

Scopus:262

No limitation of time and language

TITLE-ABS-KEY(

("multiple sclerosis" OR MS OR"demyelinating disease*" OR "demyelination disorder*")

AND

(

"deep brain stimulation"

OR DBS

OR neuromodulation

OR "brain stimulation"

OR "stereotactic stimulation"

OR "implantableneurostimulator"

OR "thalamic stimulation"

OR "VIM stimulation"

OR "ventral intermediate nucleusstimulation"

)

AND

(

tremor

OR "intention tremor"

OR "postural tremor"

OR "action tremor"

OR "MS tremor"

OR "cerebellar tremor"

))

_______________________________________

Web of science :261

No limitation of time and language

TS=(

("multiple sclerosis" OR MS OR"demyelinating disease*" OR "demyelination disorder*")

AND

(

"deep brain stimulation"

OR DBS

OR neuromodulation

OR "brain stimulation"

OR "stereotactic stimulation"

OR "implantableneurostimulator"

OR "thalamic stimulation"

OR "VIM stimulation"

OR "ventral intermediate nucleusstimulation"

)

AND

(

tremor

OR "intention tremor"

OR "postural tremor"

OR "action tremor"

OR "MS tremor"

OR "cerebellar tremor"

))

______________________________________________

Embase:301

No limitation of time and language

('multiple sclerosis':ti,ab,kw OR MS:ti,ab,kw

OR 'demyelinating disease*':ti,ab,kw

OR 'demyelination disorder*':ti,ab,kw)

AND

('deep brainstimulation':ti,ab,kw

OR DBS:ti,ab,kw

OR neuromodulation:ti,ab,kw

OR 'brain stimulation':ti,ab,kw

OR 'stereotactic stimulation':ti,ab,kw

OR 'implantable neurostimulator':ti,ab,kw

OR 'thalamic stimulation':ti,ab,kw

OR 'VIM stimulation':ti,ab,kw

OR 'ventral intermediate nucleusstimulation':ti,ab,kw)AN

D(t

remor:ti,ab,kw O

R 'intention tremor':ti,ab,kw O

R 'postural tremor':ti,ab,kw O

R 'action tremor':ti,ab,kw O

R 'MS tremor':ti,ab,kw O

R'cerebellar tremor':ti,ab,kw)
