## Supplementary material for "Therapeutic Efficacy and Safety of Deep Brain Stimulation for Multiple Sclerosis Related-Tremor: A Systematic Review and Meta-Analysis": prisma flowchart

Studies from databases/registers **(n = 1012)**

Embase (n = 301)

Scopus (n = 262)

Web of Science (n = 261)

PubMed (n = 188)

**Identification**

Studies included in review **(n = 13)**

**3 RCTs**

**1 Non-randomized trial**

**6 Retrospective cohort**

**3 Prospective cohort**

Studies excluded **(n = 464)**

Studies not retrieved **(n = 0)**

Studies assessed for eligibility **(n = 78)**

Studies sought for retrieval **(n = 78)**

Studies screened **(n = 542)**

References removed **(n = 470)**

Duplicates identified manually (n = 22)

Duplicates identified by Covidence (n = 448)

Marked as ineligible by automation tools (n = 0)

Other reasons (n = )

**Screening**

Studies excluded **(n = 65)**

conference (n = 25)

case report (n = 6)

case series (n = 17)

protocol only (n = 3)

Not related intervention (n = 2)

Not related patient population (n = 6)

Tremor outcome not extractable (n = 3)

Not related scientific question (n = 3)

Included studies ongoing **(n = 0)**

Studies awaiting classification **(n = 0)**
